# Rapid clinical diagnosis and treatment of common, undetected, and uncultivable bloodstream infections using metagenomic sequencing from routine blood cultures with Oxford Nanopore

**DOI:** 10.1101/2025.01.08.25320182

**Authors:** Kumeren N. Govender, Teresa L Street, Nicholas D Sanderson, Laura Leach, Marcus Morgan, David W. Eyre

## Abstract

**Background:** Metagenomic sequencing has the potential to transform clinical microbiology by enabling rapid pathogen identification and antimicrobial resistance (AMR) prediction in critically ill patients with bloodstream infections (BSIs). However, its clinical implementation has been hindered by challenges in speed, accuracy, and technical feasibility. We present a direct from positive blood culture workflow using Oxford Nanopore sequencing that overcomes these limitations, delivering results rapidly and accurately.

**Methods:** We developed and evaluated a direct-from-blood culture metagenomic sequencing method for rapid pathogen and AMR prediction using Oxford Nanopore Sequencing. Species prediction was performed using Kraken2 with a comprehensive standard database, employing heuristic and random forest classification models. Additionally, we benchmarked AMR classification tools and databases, including ResFinder, CARD, and NCBI AMRFinderPlus. We processed 273 randomly selected blood cultures (211 positive, 62 negative) from hospitalised patients in real time, comparing species identification, AMR detection and time-to-result against standard culture-based diagnostics performed by the routine microbiology laboratory.

**Findings:** Our method achieved 97% sensitivity and 94% specificity (both improving to 100% after accounting for plausible additional infections) for species identification compared to established diagnostic methods. We detected 18 additional infections—13 polymicrobial and 5 previously unidentifiable—and delivered findings in 3.5 hours, nearly a third of time taken by routine methods. For the top ten pathogens, our method produced AMR results 20 hours faster than current antimicrobial susceptibility testing, with 88% sensitivity and 93% specificity. For *Staphylococcus aureus* and *Escherichia coli*, AMR prediction sensitivity was 100% and 91%, and specificity 99% and 94% respectively.

**Interpretation:** These findings highlight the potential of metagenomic sequencing to improve BSI diagnosis by providing rapid and comprehensive pathogen and AMR detection. Integrating this approach into routine clinical workflows could bridge critical diagnostic gaps in sepsis care, reduce empirical antibiotic use, and inform targeted treatment within hours rather than days.

**Funding:** National Institute for Health Research (NIHR) Biomedical Research Centre, Oxford.

**Research in Context:** *Evidence before this study:* Bloodstream infections (BSIs) remain a critical global health burden, affecting over 30 million people annually with mortality rates reaching up to 30%. The urgency of early diagnosis is well-established -delays in appropriate antimicrobial therapy, even by one hour, can significantly reduce survival in severe cases. Conventional blood culture, the current diagnostic standard, is hampered by long turnaround times (24–72 hours), reduced sensitivity in patients already on antibiotics, and its inability to detect fastidious, slow-growing, or unculturable organisms. To explore current advancements in the field, we conducted a search on PubMed and Google Scholar on 6th April 2025 using a combination of relevant medical subject heading terms: “infection,” “diagnosis,” “metagenomic,” and “bloodstream.” This search yielded 59 results, from which we excluded pilot studies, those with fewer than 50 samples and studies focused on diagnosing specific pathogens. After applying these exclusions, we found four that contributed valuable insights into the application of metagenomics in BSIs. Blauwkamp and colleagues demonstrated strong concordance between cell-free DNA sequencing and blood culture, although the technique showed limited utility in predicting antimicrobial resistance. Similarly, Rossoff and colleagues used a cell-free DNA assay at a CLIA-certified laboratory demonstrating performance with >90% with turnaround times within 48 hours. Anson and colleagues developed a method to extract bacterial DNA from positive blood cultures for whole-genome sequencing, successfully identifying *Staphylococcus* and Gram-negative species, though antimicrobial resistance prediction was limited with Illumina data alone. A more recent study by Harris and colleagues demonstrated that direct-from-blood culture methods can accurately identify causative pathogens at the species level in ICU patients, particularly in monomicrobial infections, approximately 9-17 hours after a positive blood culture result. To date, no study, to our knowledge, at scale has systematically evaluated the diagnostic yield, speed, and practical clinical utility of metagenomic sequencing applied directly to randomised routine blood culture samples.

*Added value of this study:* This study presents the first clinical implementation of metagenomic sequencing using Oxford Nanopore technology directly on routine blood cultures at scale, with a focus on real-time diagnosis of bloodstream infections. It uniquely assesses the ability of metagenomics to detect not only common but polymicrobial, uncultivable, or previously undetected pathogens, and to predict antimicrobial resistance within clinically actionable timeframes. The study advances the field by demonstrating how accurate and rapid pathogen identification and resistance profiling can be integrated into existing diagnostic workflows, thereby reducing diagnostic delay and enabling early, targeted antimicrobial therapy.

*Implications of all the available evidence:* The collective evidence indicates that clinical metagenomics has reached a stage of maturity where we now see workflows ready for implementation in clinical settings. This study adds to the growing recognition that, when applied to positive blood cultures, metagenomic sequencing can bridge critical diagnostic gaps in sepsis care, reduce empirical antibiotic use, and inform precision treatment within hours rather than days. Nonetheless, widespread clinical adoption will require further standardisation of laboratory protocols, development of robust interpretive frameworks, and evidence of cost-effectiveness in real-world settings. As antimicrobial resistance continues to rise globally, metagenomics offers a robust and strategic approach to meet one of the most pressing challenges in infectious diseases.

## Introduction

Bloodstream infections (BSIs) represent a significant global health concern, affecting over 49 million individuals annually and leading to approximately 11 million sepsis-related deaths.^1^ Early diagnosis and rapid administration of appropriate antibiotic therapy is crucial; delayed or inactive antibiotics lead to significant increases in mortality rates.^2^ However, empirical antibiotic treatment can be ineffective in 20-34% of cases, with emerging antimicrobial resistance (AMR) exacerbating this problem.^3–5^ Blood culture, first described in the mid 20^th^ century, and widely automated, but largely unchanged, since the 1970s, is recognised as the standard of care for accurately identifying the causative pathogen and its antimicrobial susceptibility, essential for ensuring targeted and effective antibiotic treatment. However, blood cultures are negative in up to two thirds of probable sepsis cases due to a variety of factors including low numbers of organisms in the bloodstream, pre-exposure to antibiotics, non-infectious inflammatory disorders, or fastidious or atypical organisms.^6,7^ Moreover, blood culture is slow relative to the pace of illness in severe infections; it requires 24-72 hours to yield results, whereas in severe sepsis each hour delay in appropriate antibiotic administration may increase in mortality by 8%.^8,9^

Molecular methods offer a promising solution to the limitations of traditional culture techniques.^10^ Although multiplex PCR panels are fast, achieving results within 1-4 hours from positive blood cultures, this technology is limited by using a finite array of gene markers limiting the breath of species and resistance markers identifiable, which must also be updated as new resistance emerges.^11–14^ Most platforms start with positive blood cultures; with only one FDA-approved platform that detects infection directly from patient blood samples (T2 Biosystems), detecting 6 bacterial pathogens in 3-5 hours.^15^

In contrast, clinical metagenomics, an unbiased, sequencing-based method capable of detecting nearly all organisms within a sample, potentially offers a more comprehensive diagnostic tool.^16^ It overcomes the limitations of multiplex PCR by not relying on predefined gene markers, thereby broadening the spectrum of detectable pathogens and resistance mechanisms.^17^

Methods for sequencing bacterial DNA direct from positive blood cultures on Illumina MiSeq and Nanopore MinION systems have been previously described^18^, identifying specific Gram-negative and *Staphylococcus sp.* species and limited AMR-encoding genes within 4 hours in one study^19^ and accurate monomicrobial species prediction within 9-17 hours in another.^20^ However, such studies are relatively underpowered, limited to spiked samples or enriched for specific organisms, or have reported only AMR gene detection without assessment against phenotypic resistance. Another approach, using microbial cell-free DNA (cfDNA) sequencing, performed directly from blood samples without needing any culture, can identify a diverse array of species but needs costly high-throughput sequencing per sample, often yielding <0.01% of pathogen reads and has limited AMR predictivity.^21,22^

Here, we present an optimized, unified sample-to-result metagenomic pipeline employing Nanopore sequencing directly from positive blood cultures, using an unbiased sample selection to evaluate species identification, resistance detection, and time-to-result against a routine clinical microbiology laboratory in Oxford, UK, in real-time.

## Methods

### Sample Collection and Processing

Between November 2020 and November 2022, 211 positive and 62 negative blood cultures were processed from Oxford University Hospitals. Six random samples were typically chosen daily, excluding those collected in BD BACTEC™ Peds Plus™ media. This study was done with approval from the London Queen Square Research Ethics Committee (REC reference 17/LO/1420). Samples, collected as part of routine clinical care, were processed immediately after routine diagnostic workup. Only one bottle per culture set (aerobic or anaerobic) was analysed, using BD BACTEC™ FX systems, which incubates bottles at 35°C until flagged positive or for five days. A 1.5 mL aliquot of the sample was used for downstream processing. Additionally, colony forming units were quantified using various agar plates (See Supplementary Methods).

### DNA Extraction from Blood Culture

DNA was extracted without pre-treatment using the BiOstic Bacteraemia kit (MoBio, Qiagen, USA), with minor adjustments to the manufacturer’s protocol (Figure S1). Each batch included six clinical samples, two negative controls (pure water & uninoculated broth), and one positive control (*Arcanobacterium haemolyticum*). DNA was normalised, with internal control *Thermus thermophilus* DNA spiked into each sample (2% of the DNA concentration).

### Library Preparation and Nanopore Sequencing

DNA libraries were prepared using the Rapid Sequencing Kit with barcoding (SQK-RBK004) and sequenced on the GridION platform with R9.4.1 flow cells for 24 hours according to the manufacturer’s protocol. Each batch was processed with 6 samples and 3 controls. Live basecalling was performed with Guppy (version 3.2.6).

### Bioinformatics

#### Species prediction

Reads were demultiplexed (Guppy version 6.4.8), and low quality and length <1000bp was removed (prinseq-lite v0.20.4; Figure S2).^23^ Three methods were benchmarked for species identification using Kraken2 V2.0.7^24^ and Bracken V2.5^24^ using a full standard database (NCBI RefSeq February 13th, 2022). The first method removed plasmid reads and used read count and percentage thresholds. The second included plasmid reads and aligned reads to reference genomes using Minimap2 v2.24-r1122 and Samtools v1.17, calculating additional metrics such as coverage breadth and depth, and percentage of reads mapped from extracted species reads. Heuristic thresholds were optimised using Youden’s Index. The third method utilized a random forest model based on the same five metrics. The dataset was split 60:40 into training and test sets. Thresholds were re-optimized after correcting false positives and negatives. Reports were adjusted for bioinformatic contamination using thresholds set from simulation studies (contamination thresholds available in Table S2; method as previously described).^23^ Genotypic and phenotypic discrepancies were assessed using BLAST, genome coverage, average nucleotide identity (ANI), microbiological data, and API 20E testing.

### Antimicrobial Resistance Prediction

Genotypic resistance was assessed using ResFinder 4.0 (db downloaded July 3^rd^, 2022), Resistance Gene Identifier (CARD/mCARD downloaded August 18^th^, 2022), and NCBI AMRFinderPlus 3.10.40 (db downloaded July 30^th^, 2022). Predictions were benchmarked against phenotypic susceptibility data (BD Phoenix microbroth dilution (BD, USA)) from pure cultured isolates. We interpreted resistance at the class level given only ResFinder could predict at the drug level. A sample was considered to be genotypically sensitive if no resistant genes were found with a genome sequencing depth of at least 10x at the end of sequencing. Results were classified by categorical agreement, very major errors (VME), and major errors (ME).

### Barcode Contamination Assessment

Sequencing involved multiplexing with barcodes. Mis-assigned and cross-assigned reads were quantified using the unused barcode 3 and *Arcanobacterium haemolyticum* as a proxy. Mis-assignment rates were estimated from reads in unused barcodes, while cross-assignment was inferred from detection of *Arcanobacterium haemolyticum* in barcodes other than the positive control.

## Results

### Sample characteristics

We performed metagenomic sequencing using the Oxford Nanopore platform in 273 randomly-selected blood culture bottles (Figure 1). Of 211 positive blood cultures, 205 contained 216 species identified by the routine laboratory using MALDI-TOF (Bruker, USA), including 11 polymicrobial infections (species and antimicrobial susceptibilities listed in Table 1, Table S1). Routine methods failed to identify any species in the remaining 6 positive blood cultures. Details on negative, internal, and positive controls, and barcode demultiplexing performance are provided in the Supplementary Results.

**Figure 1.**
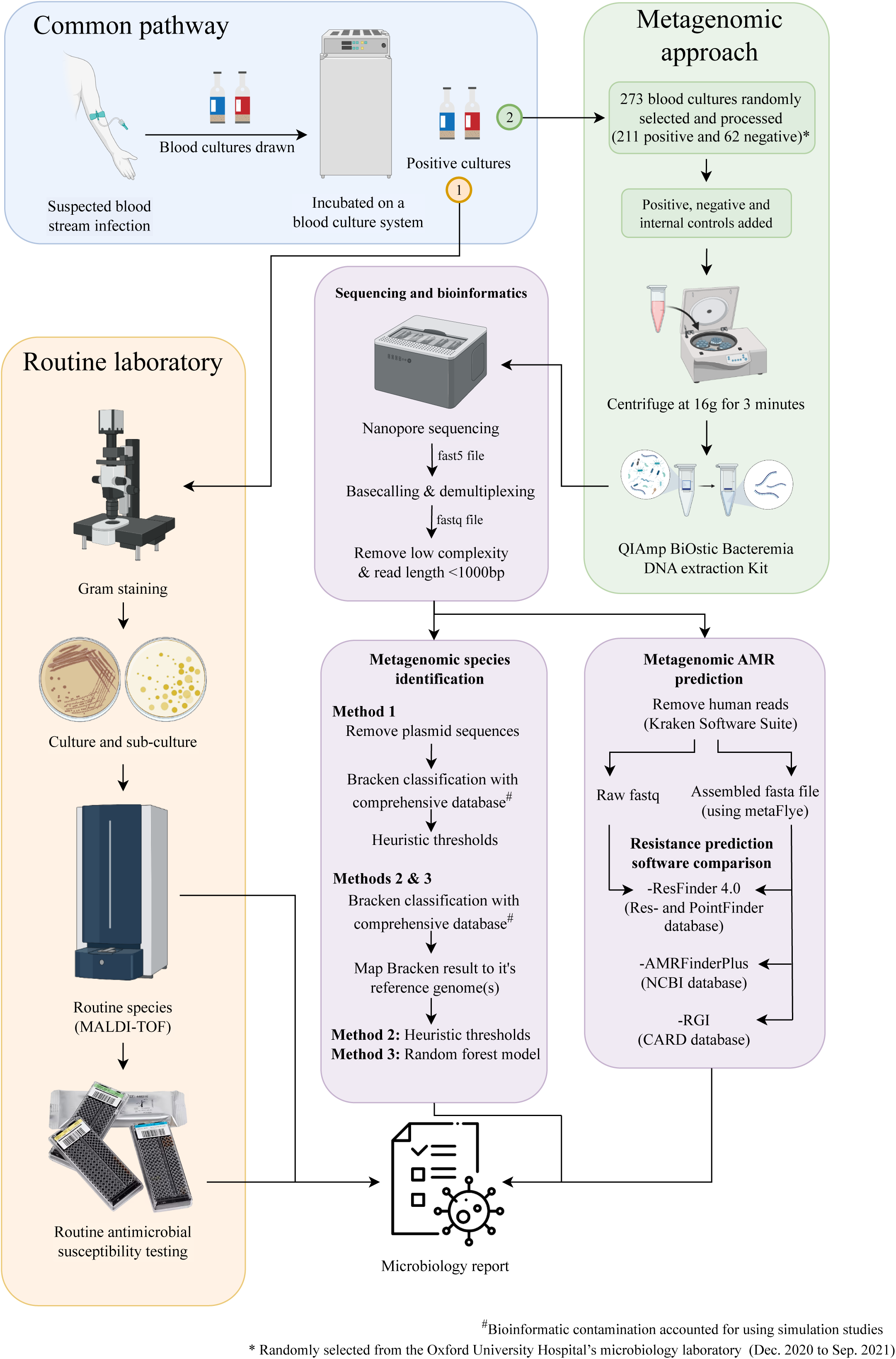
Routine microbiology laboratory vs. metagenomic study workflow. Blood cultures are drawn from patients with suspected blood stream infections and are incubated until culture positivity or negativity (defined as two weeks of no positivity). **1.** In the routine laboratory positive blood cultures undergo immediate gram-staining and are re-cultured and then often sub-cultured on a petri dish to obtain a pure culture. MALDI-ToF and antimicrobial susceptivity testing is then performed. **2.** In the metagenomic workflow, DNA is extracted directly from positive blood cultures for processing using the QIAmp BiOstic Bacteremia kit. In this study, 211 positive and 62 negative blood cultures were selected at random and processed with added positive, negative and internal controls. Sequencing was performed in batches of 6 samples per flow cell using the Oxford Nanopore GridION platform. Three bespoke methods to determine metagenomic species and three resistance prediction software tools were evaluated against routine phenotypic results.

**Table 1.**
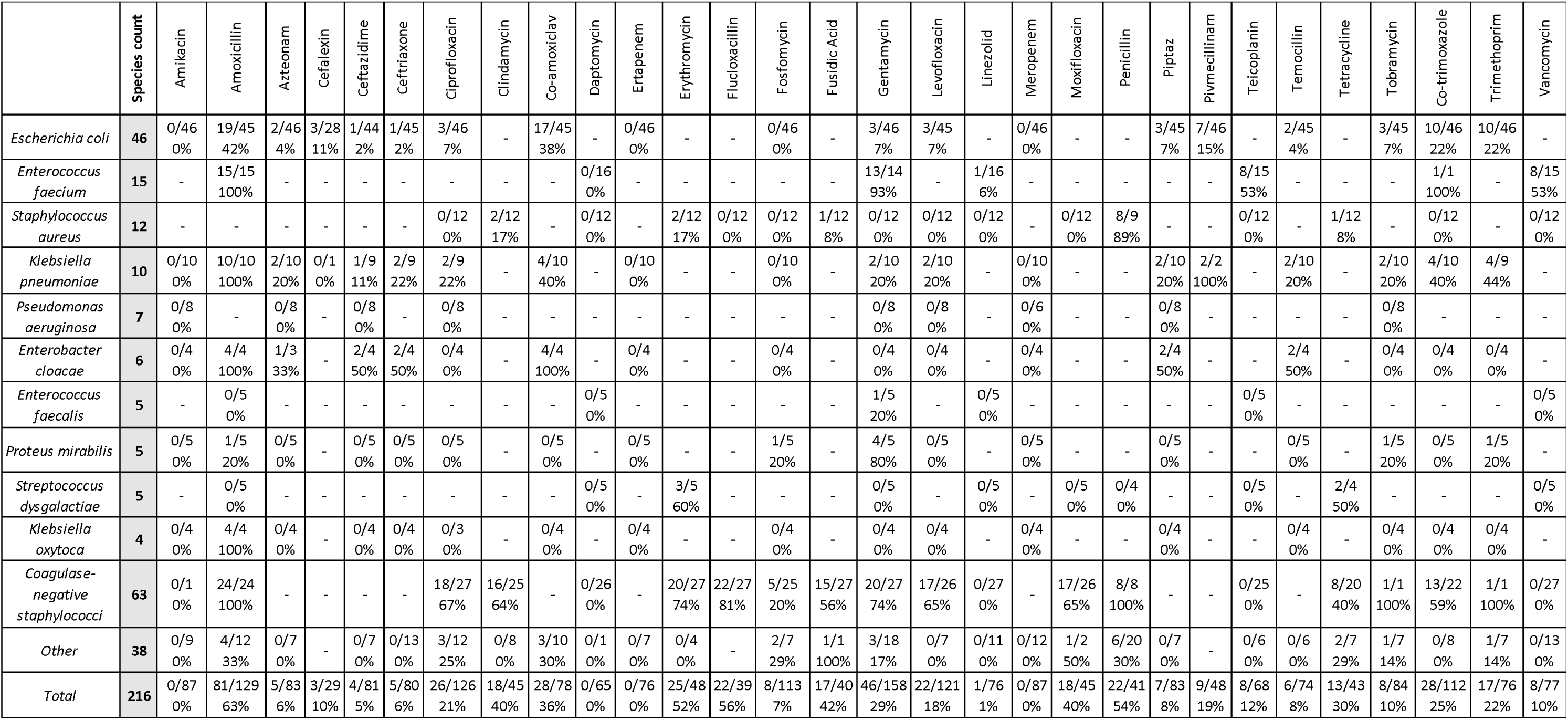
Species and resistance phenotypic summary for top 10 species. The top 10 species are displayed with phenotypic resistance data shown by number of resistant isolates/(resistant + sensitive isolates) grouped by drug. See Table S1 for further details.

### Species identification

#### Unadjusted species performance

To identify species, three bioinformatic methods using Bracken with a standard database adjusted for contamination (see Methods; thresholds in Table S2) were tested. Method 1 excluded plasmid sequences, Method 2 added genome mapping and heuristic thresholds (number of species reads and percentage of bacterial reads from the species), and Method 3 used mapping with a random forest model-derived thresholds. Metagenomics species identification was compared to routine MALDI-TOF analysis, representing “naïve” performance. Sensitivity was defined on a per species basis, i.e., the proportion of all species found by MALDI-TOF across all samples also detected by sequencing. Specificity was calculated within the positive samples, as the proportion of samples with ≥1 extra species detected by sequencing. Sensitivity and specificity in the testing datasets for Methods 1, 2 and 3 were 92% (95%CI, 85-97) and 95% (88-99), 94% (87-98) and 94% (86-98), and 92% (84-97) and 91% (83-96) respectively (Table 2, thresholds available in Table S7). Method 2 was used for subsequent analyses.

**Table 2.** Species performance results across all models. This table summarises results of models across all classification approaches used. Samples were randomly split into training (60%) and testing (40%) sets to ensure independent validation of species prediction approaches. Three different methods were used for bioinformatic species identification, all using the classification software Bracken with a standard database and adjusted for bioinformatic contamination using thresholds set from simulation studies.

|  | Training |  | Testing |  | Training & testing dataset |  |  |
| --- | --- | --- | --- | --- | --- | --- | --- |
|  | Culture positive |  | Culture positive |  | Culture positive |  | Culture negative |
|  | Sensitivity (95% CI) [sample #] | Specificity (95% CI) [sample #] | Sensitivity (95% CI) [sample #] | Specificity (95% CI) [sample #] | Sensitivity (95% CI) [sample #] | Specificity (95% CI) [sample #] | Specificity (95% CI) [sample #] |
| Plasmid removal (Method 1): naive | 97% (92-99) [124/128] | 98% (93-99) [120/123] | 92% (85-97) [81/88] | 95% (88-99) [78/82] | 94% (91-97) [201/216] | 97% (93-99) [198/205] | 98% (91-100%) [61/62] |
| Mapping and heuristic thresholds (Method 2): naive | 99% (96-100) [127/128] | 96% (91-99) [118/123] | 94% (87-98) [83/88] | 94% (86-98) [77/82] | 97% (94-99) [210/216] | 94% (90-97) [193/205] | 98% (91-100%) [61/62] |
| Mapping and random forest model (Method 3): naive | 100% (97-100) [128/128] | 100% (97-100) [123/123] | 92% (84-97%) [81/88] | 91% (83-96%) [75/82] | 97% (93-99%) [209/216] | 97% (93-99%) [198/205] | 98% (91-100%) [61/62] |
| Mapping and heuristic thresholds (Method 2): adjusted | 100% (97-100) [137/137] | 100% (97-100) [123/123] | 100% (96-100) [92/92] | 100% (97-100) [82/82] | 100% (95 CI, 98-100) [229/229] | 100% (95 CI, 98-100) [205/205] | 100% (94-100%) [61/61] |
| Mapping and random forest model (Method 3): adjusted | 100% (97-100) [137/137] | 100% (97-100) [123/123] | 100% (96-100) [92/92] | 100% (96-100) [82/82] | 100% (95 CI, 98-100) [229/229] | 100% (95 CI, 98-100) [205/205] | 100% (94-100%) [61/61] |

\*Spe
cificity was calculated on a per sample basis, where any sample with >1 additional species found from sequencing was considered to contribute as 1 to the false negative count.

#### False-negative results (Method 2)

Using Method 2, within both training and test datasets, there were 6 false-negative results where sequencing failed to detect a species found by MALDI-TOF. In four, sequencing identified a species similar to MALDI-TOF that was potentially a more accurate representation of the species (Table S3, Methods). The other two missed species (samples 143 and 147) contained reads from the species, but not sufficient to pass thresholds set for calling these samples positive; of note both samples were polymicrobial by culture.

We therefore investigated relaxing the stringency of filtering to optimise sensitivity, selecting new species identification thresholds (50 species reads, 0.5% of all bacterial reads) that achieved 100% sensitivity in training data while also maximising specificity (Figure S6). Using these thresholds, sensitivity and specificity in the test dataset were 100% (95% CI, 97-100%) [128/128] and 94% (88-97%) [115/123] respectively, and 98% (95-99%) [212/216] and 92% (88-95%) [189/205] in the whole dataset. The four remaining false-negative results were explained by MALDI-TOF/sequencing discrepancies described above.

#### False-positive results (Method 2)

Additional species detected by sequencing but not by routine culture and MALDI-TOF, were assessed for bioinformatic plausibility (see Methods). 16 samples had false-positive species on sequencing, including the four MALDI-TOF/sequencing discrepancies described earlier as also false-negatives. In the 12 other samples, 13 additional species were found using sensitivity-optimised thresholds (Table 3). These were considered biologically plausible and likely true infections (Table 3; Table S4). Mapping was used to confirm the taxonomic classification, 11/13 species had >75% genome coverage and depths between >3x and 143x. The other two species, *Collinsella aerofaciens* (48% coverage, 3x depth) and *Bacteroides nordii* (68% coverage, 1.5x depth alongside *Bacteroides fragilis* and *Bacteroides thetaiotaomicron*) had lower sequence coverage, but could plausibly have been part of polymicrobial infections although could also represent difficulties classifying genetically similar species for *Bacteroides*.^25^ Anaerobes were over-represented compared to their prevalence in cultures (5/13 species), suggesting limitations in current microbiological methods for detecting such organisms, e.g. *Bacteroides fragilis* was linked to liver abscess in sample 78.

**Table 3.**
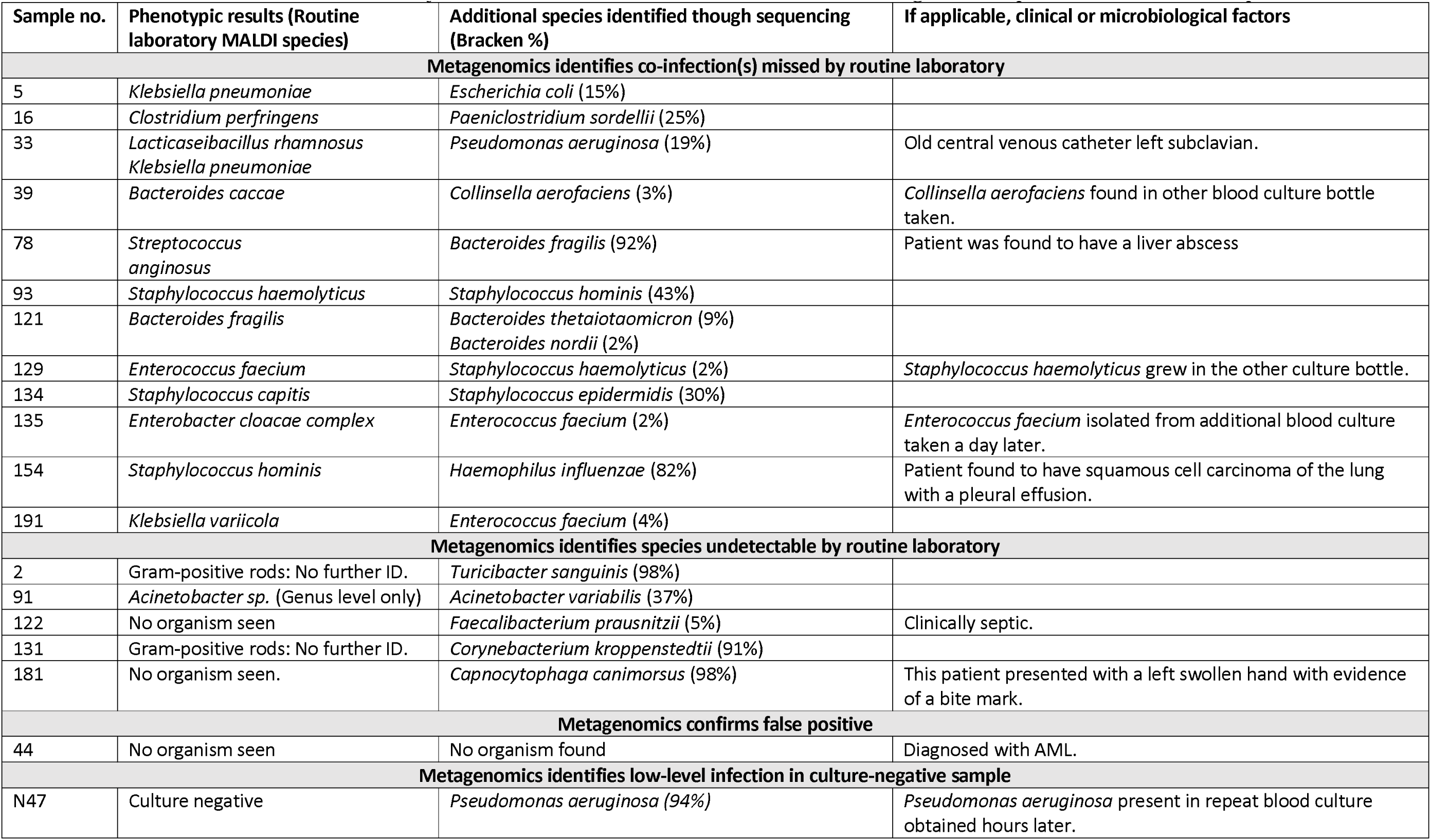
Species missed by routine phenotypic methods identified by sequencing pipeline. Metagenomic sequencing identifies missed co-infections, traditionally unidentifiable, low-level infections in culture-negative samples and confirms false positives.

None of the additional species had high ANIs (>95%) with those detected phenotypically, indicating their distinct nature. In samples 33, 39, and 129, low levels of *Pseudomonas aeruginosa*, *Collinsella aerofaciens*, and *Staphylococcus haemolyticus* were identified respectively, which routine methods missed in initial cultures but detected in subsequent tests. This suggests some organisms present at low levels in blood cultures may evade standard microbiological detection.

#### Reviewing thresholds accounting for plausible additional species

As we considered most or all of the additional species identified as plausible, we also tested further lowering thresholds for species detection (50 reads, 0.1%), selecting the lowest level that still accounted for most contamination in simulations. This adjustment resulted in 100% sensitivity and 99.5% specificity until a 0.4% cut-off, where both improved to 100% (Figure S7). A single false positive, likely due to *Staphylococcus aureus* bioinformatic misclassification with poor genome coverage, impacted lower-threshold performance (Table S6). Using these final filtering thresholds, sensitivity and specificity were both 100% in both the training and testing datasets (95%CI 97-100% and 96-100% respectively) (Table 2).

#### False negative and false positive results (Method 3)

Results using Method 3 were similar. Before adjustment for plausible species, the seven false negative results included the 4 MALDI-TOF misclassifications previously described (Table S3), and 3 other missed species (Table S5). Within the seven false positive results in the testing data, four were likely due to MALDI-TOF inaccuracy (Table S3) and three had been previously assessed as being plausible. This approach failed to identify a result that was considered plausible in the threshold-based analysis (Table S5). The total number of reads was the most important feature in the random forest model trained with adjusted plausible species (Figure S5B). After retraining models accounting for plausible additional species, sensitivity and specificity were both 100% in both the training and testing datasets (95%CI 97-100% and 96-100% respectively), matching Method 2.

#### Culture negative samples

In all methods, overall unadjusted species-level specificity in culture-negative samples was 61/62 (98%; 95%CI 91-100%), and after adjusting for a plausible infection was 61/61 (100%; 94-100%) (Table 2). One sample, Sample N47 was negative by the routine laboratory, however, sequencing confirmed the presence of *Pseudomonas aeruginosa*, which was also present in repeat blood culture obtained 24 hours later from the same patient. The number of bacterial reads before filtering was otherwise consistently low (Figure S8).

#### Positive blood cultures unidentifiable by the routine lab

Six blood cultures flagged positive, indicating potential bacterial growth, but species identification was not achieved using routine techniques (Table 3). Sample 44 likely represents a false positive ‘flag’ due to the patient’s acute myeloid leukemia, where highly active white blood cells may have triggered the flag. MALDI-TOF provided only genus-level identification for sample 91, but sequencing identified *Acinetobacter variabilis*. Notably, sequencing uncovered organisms missed by conventional methods in samples 2, 122, 131, and 181, identifying them as *Turicibacter sanguinis*, *Faecalibacterium prausnitzii*, *Corynebacterium kroppenstedtii*, and *Capnocytophaga canimorsus*, respectively.

#### Time to species identification

We applied the threshold filtering method derived from the training dataset after sequencing ended (24 hours) to earlier sequencing time intervals. Analysing data available at 10 minutes after sequencing started resulted in a sensitivity and specificity of 78% (95CI% 72-83) [178/229] and 100% (98-100) [205/205] respectively, while sensitivity improved to sequencing end (Figure 2A). Using dynamic filtering thresholds based on mapping and the time since sequencing started (Table S8), sensitivity and specificity were 98% (95-99) [224/229] and 100% (98-100) [205/205] respectively 10 minutes after starting sequencing. This improved to 100% (98-100) [229/229] and >99% (97->99%) [204/205] after 120 minutes, even in the presence of polymicrobial infections with a low read abundance of 2% (Figure 2B).

**Figure 2.**
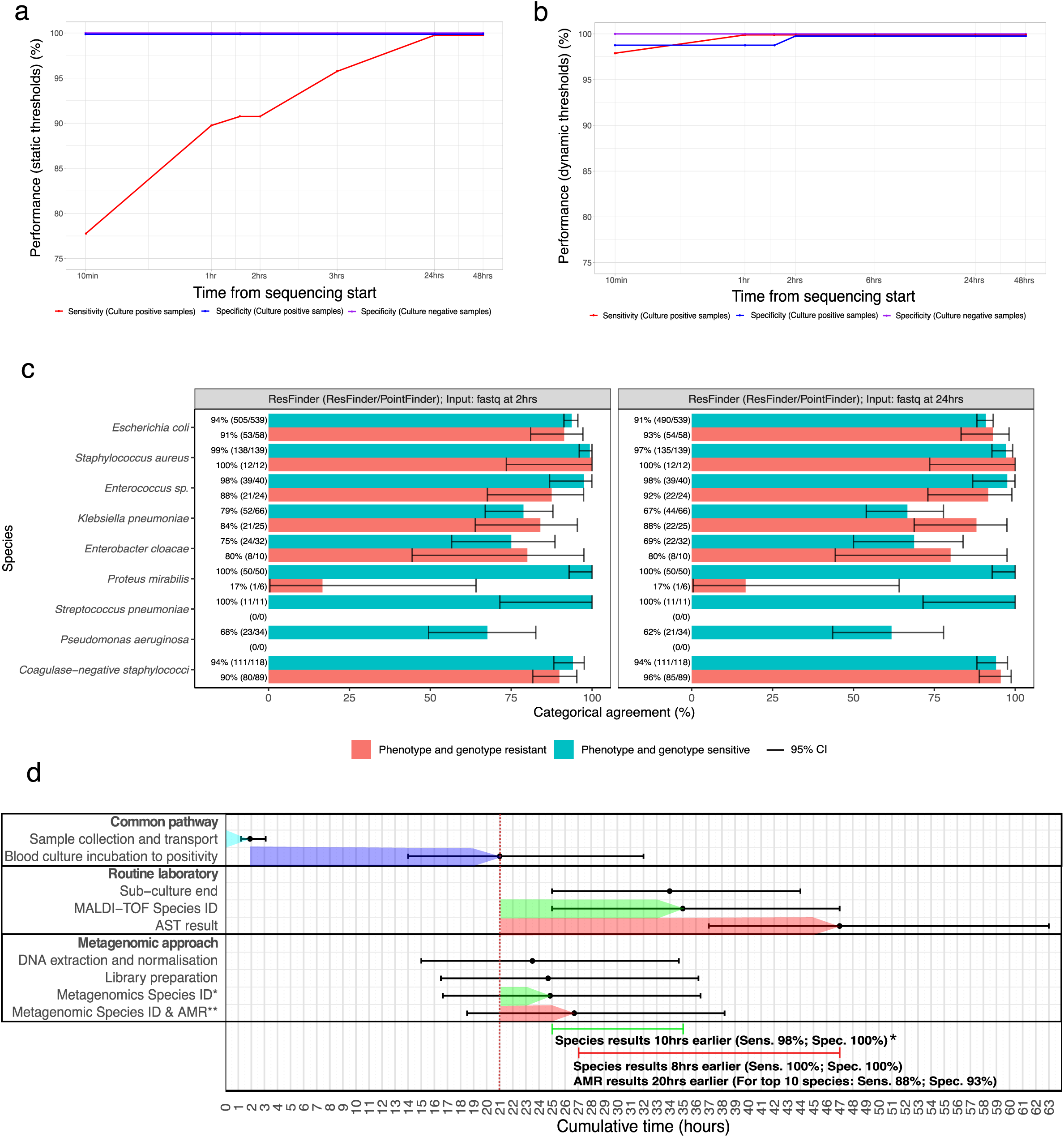
Time to species identification and antimicrobial resistance prediction from metagenomics vs. routine microbiology lab. **A.** Species identification performance by time with a static threshold applied. **B.** Species identification performance by time with a dynamic threshold applied (Table S8). **C.** Top AMR performing tools at 2- and 24-hour sequencing intervals for top 10 species. AMR tools predicting resistance to isolates were categorically evaluated against phenotypic antimicrobial susceptibility testing (AST) results obtained from the BD Phoenix microbroth dilution (BD, USA) using pure isolates. Results that were determined to be “intermediate” were excluded from the analysis. **D.** Time to metagenomic species and AMR results vs. routine microbiology laboratory.

### Antimicrobial resistance prediction

Amongst the ten most frequently isolated species, phenotypic resistance was present in 18% (224 resistant, 1029 sensitive) of drug-organism combinations in monomicrobial samples (Table 1). The predictive capacity of drug resistance was evaluated through a comparative analysis with four software and database tools in these top ten species.

Using assembled genomes from data generated by 24hrs of sequencing, ResFinder had the highest overall sensitivity and specificity of 84% (95%CI 78-88%, 188/224) and 96% (94-97%, 983/1029) respectively, while mCARD had the lowest, 76% (70-81%, 170/224) and 76% (74-79%, 786/1029) respectively (Table 4).

**Table 4.** Genotypic performance of top 10 clinically relevant species by tool. Four AMR prediction tools were benchmarked including ResFinder, AMRFinderPlus, Resistance Gene Identifier (RGI), and RGI with a modified database. Resistance was called from genome assembly, raw reads from the end of the sequencing run and raw reads at earlier sequencing time intervals.

| Database | Software | Input format | Sequencing start time to result | Resistant agreement | Sensitive agreement | Major Error rate | Very Major Error rate |
| --- | --- | --- | --- | --- | --- | --- | --- |
| **Resistance called after genome assembly** |  |  |  |  |  |  |  |
| ResFinder and PointFinder database | ResFinder | fasta | 24 hours | 84% (78-88%, 188/224) | 96% (94-97%, 983/1029) | 4% | 16% |
| NCBI's AMRFinderPlus database* | AMRFinderPlus | fasta | 24 hours | 79% (74-85%, 178/224) | 79% (77-82%, 816/1029) | 21% | 21% |
| Comprehensive Antibiotic Resistance Database (CARD)* | Resistance Gene Identifier (RGI) | fasta | 24 hours | 79% (73-84%, 177/224) | 77% (74-79%, 788/1029) | 23% | 21% |
| ResPipe: modified Comprehensive Antibiotic Resistance Database (CARD)* | Resistance Gene Identifier (RGI) | fasta | 24 hours | 76% (70-81%, 170/224) | 76% (74-79%, 786/1029) | 24% | 24% |
| **Resistance called using raw reads (no genome assembly)** |  |  |  |  |  |  |  |
| ResFinder and PointFinder database | ResFinder | fastq | 24 hours | 91% (87-94%, 204/224) | 90% (88-91%, 923/1029) | 10% | 9% |
| **Resistance called using raw reads at earlier time intervals (no genome assembly)** |  |  |  |  |  |  |  |
| ResFinder and PointFinder database | ResFinder | fastq | 2 hours | 88% (82-92%, 196/224) | 93% (91-94%, 953/1029) | 7% | 12% |
| ResFinder and PointFinder database | ResFinder | fastq | 1 hour | 82% (76-87%, 184/224) | 94% (92-95%, 963/1029) | 6% | 18% |

\*These tools predicted genotypic resistance at class level only, which was interpreted as resistance at all drug levels within that specific class.

Using ResFinder and raw fastq reads available after 24 hours of sequencing as input yielded a higher sensitivity and specificity, 91% (95% CI: 87-94%, 204/224) and 90% (95% CI: 88-91%, 923/1029) respectively, compared to using assembled genomes. We also investigated ResFinder sensitivity and specificity, using raw reads available after 1 hour of sequencing, 82% (95% CI: 76-87%, 184/224) and 94% (95% CI:92-95%, 963/1029), and 2 hours of sequencing, 88% (82-92%, 196/224) and 93% (91-94%, 953/1029) respectively.

### Performance by species

For *Escherichia coli,* sensitivity and specificity with ResFinder [Input fastq at 2hrs] was 91% [95%CI 80-97%] (53/58 drug-organism combinations) and 94% [92-96%] (505/539) respectively, and for *Staphylococcus aureus* was 100% [77-100%] (12/12) and 99% [96-100%] (138/139) respectively (see Figure 2C for other species).

Across species, ResFinder-based tools generally outperformed others, but RGI (CARD) showed better performance for *Klebsiella pneumoniae* and *Enterobacter cloacae*, with 100% sensitivity and 95-100% specificity, compared to 88% and 69% for ResFinder. AMRFinder also performed similarly well for these species. Sensitivity was low for *Proteus mirabilis* (17% across tools) and for coagulase-negative staphylococci (58-65% with AMRFinder and RGI). Specificity issues were present for *Pseudomonas aeruginosa* (15-74%). Sensitivity could not be assessed for *Streptococcus pneumoniae* and *Pseudomonas aeruginosa* due to no resistant cases.

### Performance by drug and drug class

ResFinder [fastq reads at 24 hrs] had the highest sensitivity across drug classes, achieving 100% for glycopeptides (12/12), amphenicols (1/1), macrolides (33/33), and steroid antibacterials (13/13), but lower for tetracyclines (83%, 5/6) and quinolones (71%, 5/7). AMRFinder and RGI (CARD) outperformed ResFinder for beta-lactams and folate pathway antagonists (Figure S9, S10, S11, see supplement for details of genes implicated in true and false calls).

#### Polymicrobial Performance

The overall sensitivity and specificity at the drug level using ResFinder [Input: fastq reads at 24hrs] for clinically relevant polymicrobial isolates was 96% (95% CI, 79-100%, 26/27) and 74% (56-89%, 26/35) respectively (Table S11). The polymicrobial sample containing *Enterococcus faecium* and *Staphylococcus haemolyticus* had the highest sensitivity and specificity performance of 100% (10/10) and 100% (4/4) respectively, while the sample containing *Escherichia coli* and *Providencia rettgeri* had the lowest, 0% (0/5) and 100% (9/9).

### Time-point analysis vs routine laboratory

Our metagenomic workflow reported species results within 3hrs 20min (IQR, 3hrs7min-3hrs27min) [Range, 2hrs45min-3hrs48min] from obtaining positive blood cultures, i.e. including time required for DNA extraction and library preparation as well as the sequencing time. At this time point, after 10 minutes of sequencing, we achieved a sensitivity and specificity of 98% (95CI%, 95-99) [224/229] and 100% (98-100) [205/205], outperforming routine methods by ≥10 hours (Figure 2D). Additionally, our method offered actionable AMR results 20 hours earlier for the top ten pathogens, with 88% sensitivity and 93% specificity.

### Cost analysis

The total consumable cost estimated per sample processed was $105 (Table S12). Multiplexing 22 samples, which would still result in a >20x coverage for a species identified, would cost $40 per sample, while multiplexing by 94, that would provide a species only result, would cost $20 (Table S13).

## Discussion

For over a century, microscopy, culture, and immunodiagnostics have been the gold standards for infection diagnosis.^26^ In this study, we present a direct-from-blood culture metagenomic workflow using Oxford Nanopore, enabling rapid pathogen identification and AMR prediction in only 3 hours and 30 minutes in a clinical setting using randomly selected samples. This approach significantly shortened the time to species identification by at least 10 hours and AMR prediction by 20 hours, while detecting 18 missed infections, including 13 undetected polymicrobial cases and 5 that were unidentifiable by routine methods.

A key finding of this study was the 100% sensitivity and specificity achieved in detecting plausible infections using metagenomic sequencing compared to standard microbiological techniques. Additional infections identified were supported by clinical, microbial, and bioinformatic evidence. This suggests metagenomic sequencing could reveal a broader spectrum of pathogens involved in bloodstream infections than currently recognised by conventional diagnostic methods, including better detecting fastidious organisms, closely-related species causing co-infections, and minority infections that are hidden behind dominant growth in cultures. For example, sequencing identified *Bacteroides fragilis* in a patient with a liver abscess that the routine laboratory missed, *Enterococcus faecium* was detected as a co-infection by sequencing but only cultured from samples taken a day later, and in another sample *Pseudomonas aeruginosa* was detected with sequencing, but only cultured later in a subsequent blood culture from the same patient. Notably, five out of the 13 additional species identified were anaerobes, suggesting that current diagnostic methods may not be fully optimised to grow and detect these organisms, and they may be a more common cause of infection than currently thought. However, whether additional information provided by metagenomics will consistently improve clinical outcomes remains to be determined. For instance, anaerobes found in bloodstream infections related to intra-abdominal infections may already be covered by empirical therapies. However, detected species such as *Capnocytophaga canimorsus* did lead to a change in antibiotic treatment during the study, underscoring the potential clinical relevance of these findings. Ultimately, randomised trials will be needed to determine whether metagenomic sequencing truly impacts patient outcomes.

Early detection of antimicrobial resistance through metagenomic sequencing, as demonstrated by the resistance patterns identified in our study, provides a critical advantage in managing bloodstream infections. Our method delivered AMR results 20 hours faster, with 88% sensitivity and 93% specificity for the top 10 pathogens. Specifically, for the common pathogens *Staphylococcus aureus* and *Escherichia coli*, sensitivity reached 100% and 91%, with specificities of 99% and 94%, respectively. Sequencing could play a key role identifying or excluding resistance, enabling faster switches to targeted antibiotics, and reducing reliance on empirical broad-spectrum agents.

Our results are consistent with previous studies demonstrating the feasibility of nanopore-based sequencing for rapid pathogen identification and resistance profiling.^19,20^ Our study builds upon this by extending the application of metagenomics to polymicrobial infections and incorporating a bioinformatic approach that significantly improves species identification and AMR prediction, even in complex clinical samples. By optimising filtering and species identification thresholds, we could enhance the detection of low-abundance species, which are often missed by traditional methods.

Despite these promising results, several challenges remain in integrating metagenomic sequencing into routine clinical practice. Contamination control, particularly in highly multiplexed workflows, remains a critical issue. Although our internal controls demonstrated minimal cross-contamination between samples, stringent filtering thresholds are required to avoid misclassification of species, especially in low-density infections. Additionally, the cost of sequencing, while rapidly decreasing, remains higher than that of traditional culture methods, particularly when performed at scale. However, multiplexing strategies, as used in this study, can reducing per-sample costs. Limitations in reference databases for species identification and resistance gene prediction can restrict performance, studies to improve their comprehensiveness are required. Future efforts should focus on sequencing prior to blood culture positivity, and on the development of superior host DNA depletion techniques, both aimed at further reducing time to result.

In conclusion, metagenomic sequencing with Oxford Nanopore offers a potentially transformative approach to diagnosing bloodstream infections. Our findings demonstrate that this technology can provide rapid, accurate, and comprehensive pathogen identification and resistance profiling, significantly outperforming traditional culture-based methods in speed and sensitivity. With further validation in larger, multicentre clinical studies, metagenomics has the potential to become a cornerstone of precision diagnostics in infectious diseases, which ultimately may lead to improved patient outcomes by enabling timely, targeted therapies.

## Supporting information

Supplementary Methods and Results

## Data Availability

All clinical sample sequence data are available via European Nucleotide Archive (ENA) under study accession number PRJEB81413.

https://www.ebi.ac.uk/ena/browser/view/PRJEB81413?show=xrefs

## Authors and contributors

All authors contributed to conceptualization, methodology, software, formal analysis, investigation, resources, data curation, and visualisation. Writing - original draft preparation: KG; writing - review and editing, supervision and funding: NS, TS, DWE.

