## Supplementary Figures for "Rapid clinical diagnosis and treatment of common, undetected, and uncultivable bloodstream infections using metagenomic sequencing from routine blood cultures with Oxford Nanopore"

**Supplementary Information**

1. Spin 1.5ml blood in a tube at max speed (16g) for 3min. Discard supernatant.
2. Add **450μl of solution MBL** to the pellet and resuspend by pipetting. Transfer the lysate into a 2ml powerbead tube and close. Vortex for 10s to mix and place in a 70°C heat block for 15min.
3. Secure the 2ml microbead tube horizontally on the Fast Prep instrument at 5.5m/s for 30 sec X1.
4. Centrifuge the 2ml MicroBead Tube to pellet debris at 10,000g for 1min. Transfer the supernatant to a new 2ml collection tube (provided).
5. Add **100μl of Solution IRS** and vortex to mix. Incubate for 5 min at room temperature.
6. Centrifuge at 10,000g for 1 min to pellet debris. Transfer the supernatant to a new 2ml collection tube (provided). Discard pellet.
7. Add **800ul** of Solution BB. Pipette or pulse vortex to mix. Briefly centrifuge to collect any liquid from the top of the lid.
8. Load **650μl of lysate** onto a Spin Column and centrifuge at 10,000g for 1min.
9. Discard the flow-through liquid and place the Spin Column back into the 2ml collection tube. Repeat until all the lysate has been loaded onto the Spin Column.
10. Transfer the Spin Column to a new 2ml collection tube (provided) and wash by adding **500μl of solution CB.** Centrifuge 10, 000g for 1 min. Discard the flow-through and put the Spin Column back into the 2ml collection tube.
11. Wash with another **500μl of solution CB** and spin at 10, 000g for 1 min. Discard the flow-through and put the Spin Column back into the 2ml collection tube.
12. Centrifuge at 13,000g for 2min to dry the Spin Column membrane.
13. Transfer the Spin Column to a new 2ml collection tube (provided).
14. Elute by adding **30μl of solution EB** directly in the center of the membrane. Allow the Spin Column to sit at room temperature for up to 5min to maximise the elution. (Do not heat elution buffer).
15. Centrifuge at 10, 000g for 1min.
16. Discard the Spin Column and cap the 2 ml collection tube containing the genomic DNA.

**Supplementary Figure 1. DNA Extraction protocol using the QIAmp BiOstic Bacteremia DNA Kit.** This figure details the DNA extraction protocol used which has been adapted for use from direct-from-blood culture samples.

A. **Guppy demultiplexing code:**

(guppy_barcoder --input_path basecalled_fastq --save_path output_fastq --barcode_kits SQK-RBK004 -q 0 -t 10 --detect_mid_strand_barcodes --min_score_barcode_front 60 --num_extra_bases_trim 50)

B. **Prinseq code to filter low quality reads, and reads <1000bp:**

(prinseq-lite.pl -fastq input_reads -lc_method dust -lc_threshold 7 -out_bad null -out_good output_reads -min_len 1000)

C. **Building kraken2 database:**

kraken2-build --db . --download-library archaea, bacteria, viral, human, plasmid

D. **Building kraken2 database (plasmid only):**

kraken2-build --db . --download-library plasmid

E. **Extracting plasmid sequences from fastq files:**

kraken2 --db kraken2_plasmid_db sample.fastq --report output.txt --unclassified-output.fastq --confidence 0.4

F. **Classifying sequences using Kraken2:**

kraken2 --db standard_13_2_22 sample.fastq --report output.txt --output standard_kraken_report.txt

G. **Converting Kraken2 report to Bracken report:**

bracken -d standard_13_2_22 -i standard_kraken_report.txt -o standard_bracken_report.txt

**Supplementary Figure 2.** **Code used for bioinformatic software tools.** A. Demultiplexing fastq files using Guppy software. B. Prinseq code to filter low quality reads and reads <1000bp. C. Code used to build kraken2 database. D. Code used to build kraken2 database (plasmid only). E. Code to extract plasmid sequences from fastq files. F. Classification of sequences using Kraken2. G. Code to convert Kraken2 report to Bracken report.

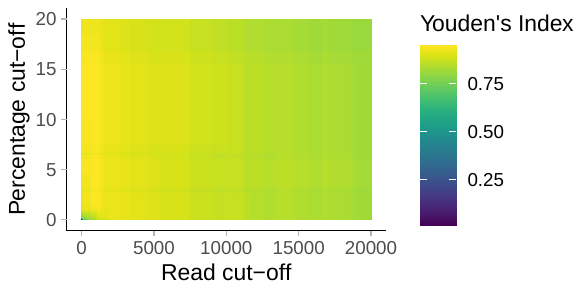
A.

B.

| **Percentage cut-off** | **Read**  **cut-off** | **Sensitivity (95% CI)** | **Specificity (95% CI)** | **Youden's Index** |
| --- | --- | --- | --- | --- |
| 10 | 750 | 97% (92-99) | 98% (93-99) | 94.4% |
| 10 | 1250 | 97% (92-99) | 98% (93-99) | 94.4% |
| 10 | 1300 | 96% (91-99) | 98% (93-99) | 93.6% |
| 10.1 | 0 | 97% (92-99) | 98% (93-99) | 94.4% |
| 10.1 | 1250 | 97% (92-99) | 98% (93-99) | 94.4% |
| 10.1 | 1300 | 96% (91-99) | 98% (93-99) | 93.6% |
| 10.1 to 15.6 | 0 to 1250 | 97% (92-99) | 98% (93-99) | 94.4% |
| 15.6 | 0 | 97% (92-99) | 98% (93-99) | 94.4% |
| 15.6 | 1250 | 97% (92-99) | 98% (93-99) | 94.4% |
| 15.6 | 1300 | 96% (91-99) | 98% (93-99) | 93.6% |

C.

| **Dataset** | **Percentage cut-off** | **Read cut-off** | **Sensitivity** | **Specificity** | **Youden's Index** |
| --- | --- | --- | --- | --- | --- |
| Training | 10 | 750 | 97% (92-99) | 98% (93-99) | 94.4% |
| Testing | 10 | 750 | 92% (85-97) | 95% (88-99) | 87.1% |
| Full set | 10 | 750 | 94% (91-97) | 97% (93-99) | 91.4% |

**Supplementary Figure 3. Deriving optimal heuristic cut-off thresholds for Method 1.** For Method 1, the optimised sensitive thresholds were 750 reads with a percentage cut-off of 10%. A. Filtering calibration heat map from training dataset. B. Method 1 applied to the training dataset: Green highlight indicates maximum Youden’s index reached. C. Method 1 applied to the training, testing and full dataset: Read and percentage cut-off maximising the Youden's Index.

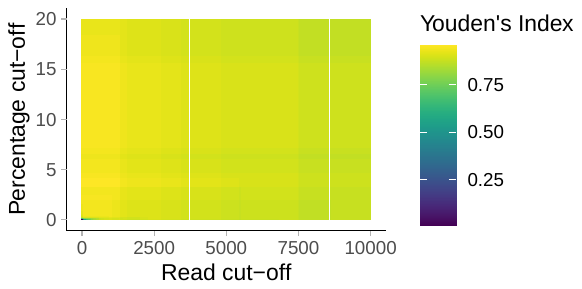
A.

B.

| **Percentage cut-off** | **Read**  **cut-off** | **Sensitivity (95% CI)** | **Specificity (95% CI)** | **Youden's Index** |
| --- | --- | --- | --- | --- |
| 3.2 | 0 | 99% (96-100) | 94% (89-98) | 93.5% |
| 3.2 | 1300 | 99% (96-100) | 94% (89-98) | 93.5% |
| 3.2 | 1350 | 98% (93-100) | 94% (89-98) | 91.9% |
| 3.3 | 0 | 99% (96-100) | 96% (91-99) | 95.1% |
| 3.3 | 1300 | 99% (96-100) | 96% (91-99) | 95.1% |
| 3.3 | 1350 | 98% (93-100) | 96% (91-99) | 93.5% |
| 3.3 to 4.1 | 0 to 1300 | 99% (96-100) | 96% (91-99) | 95.1% |
| 4.1 | 0 | 99% (96-100) | 96% (91-99) | 95.1% |
| 4.1 | 1300 | 99% (96-100) | 96% (91-99) | 95.1% |
| 4.1 | 1350 | 99% (96-100) | 96% (91-99) | 93.5% |
| 4.2 | 0 | 98% (94-100) | 96% (91-99) | 94.4% |
| 4.2 | 1300 | 98% (94-100) | 96% (91-99) | 94.4% |
| 4.2 | 1350 | 97% (92-99) | 96% (91-99) | 92.8% |

C.

| **Dataset** | **Percentage cut-off** | **Read cut-off** | **Sensitivity (95% CI)** | **Specificity (95% CI)** | **Youden's Index** |
| --- | --- | --- | --- | --- | --- |
| Training | 3.3 | 0 | 99% (96-100) | 96% (91-99) | 95.1% |
| Testing | 3.3 | 0 | 94% (87-98) | 94% (86-98) | 88.2% |
| Full set | 3.3 | 0 | 97% (94-99) | 94% (90-97) | 91.3% |

**Supplementary Figure 4. Deriving optimal heuristic cut-off thresholds for Method 2.** For Method 2, the optimised sensitive thresholds were 0 reads with a percentage cut-off of 3.3%. A. Filtering calibration heat map from training dataset. B. Method 2 applied to the training dataset: Green highlight indicates maximum Youden’s index reached. C. Method 2 applied to the training, testing and full dataset: Read and percentage cut-off maximising the Youden's Index.

A.

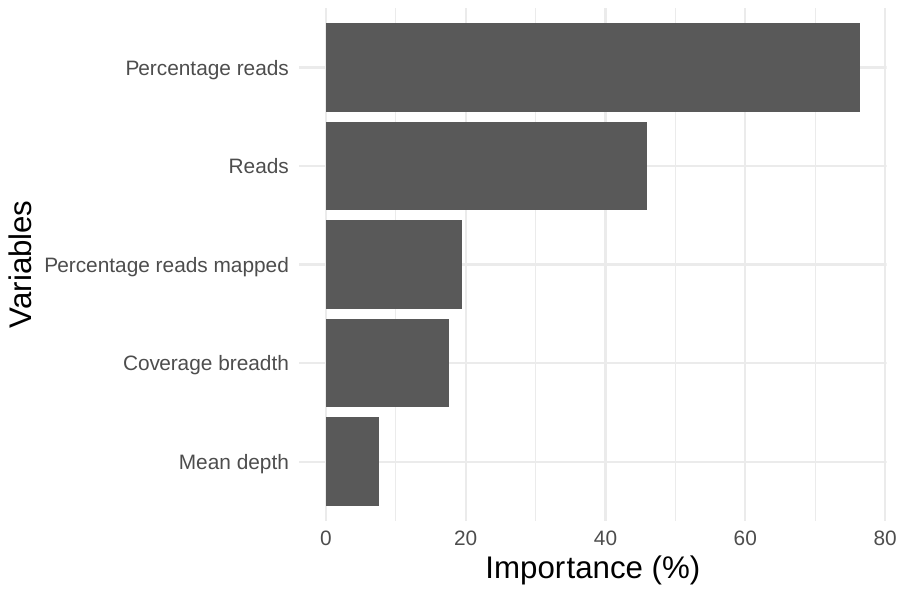

B.

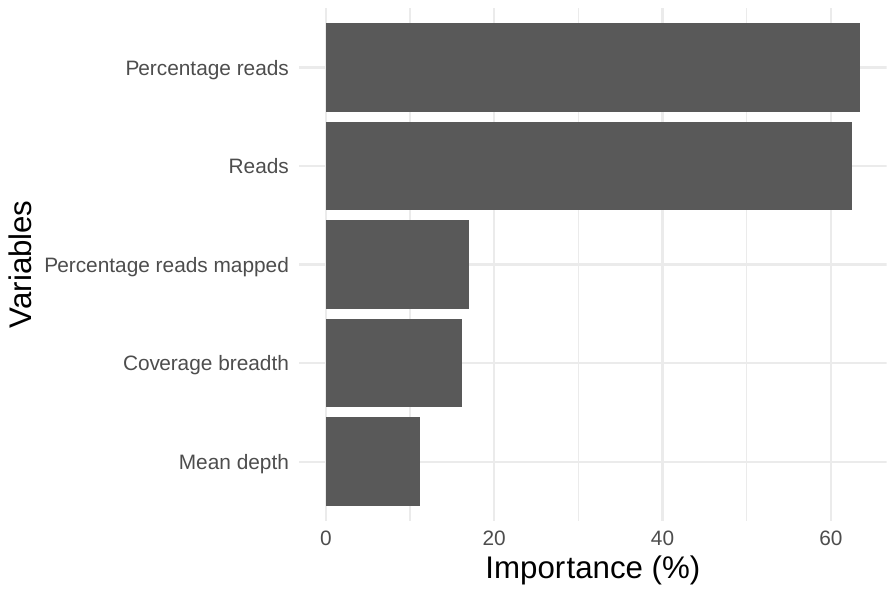

**Supplementary Figure 5. Random forest variable importance plot.** Method 3 used ranked variables. A. Random Forest variable importance plot on unadjusted performance. B. Random Forest variable importance plot on adjusted performance.

A.

| **Dataset** | **Percentage cut-off** | **Read cut-off** | **Sensitivity (95% CI)** | **Specificity (95% CI)** |
| --- | --- | --- | --- | --- |
| Training | 0.5 | 50 | 100% (97-100%) [128/128] | 94% (88-97%) [115/123] |
| Testing | 0.5 | 50 | 95% (89-99%) [84/88] | 93% (85-97%) [76/82] |
| Full set | 0.5 | 50 | 98% (95-100%) [212/216] | 92% (88-95%) [189/205] |

B.

| **Percentage**  **cut-off** | **Read**  **cut-off** | **Sensitivity**  **(95% CI)** | **Specificity**  **(95% CI)** |
| --- | --- | --- | --- |
| 0.4 | 50 | 98% (95-99%) | 92% (87-97%) |
| 0.4 | 1000 | 98% (95-99%) | 92% (87-97%) |
| 0.5 | 0 | 98% (95-99%) | 91% (86-94%) |
| 0.5 | 50 | 98% (95-99%) | 92% (88-95%) |
| 0.5 | 1000 | 98% (95-99%) | 92% (88-95%) |
| 0.5 | 1050 | 98% (95-99%) | 92% (87-97%) |
| 0.5 to 1.3 | 50 to 1000 | 98% (95-99%) | 92% (88-95%) |
| 1.2 | 0 | 98% (95-99%) | 92% (87-97%) |
| 1.3 | 50 | 98% (95-99%) | 92% (88-95%) |
| 1.3 | 1000 | 98% (95-99%) | 92% (88-95%) |
| 1.3 | 1050 | 98% (95-99%) | 92% (87-97%) |
| 1.4 | 50 | 98% (95-99%) | 92% (87-97%) |
| 1.4 | 1000 | 98% (95-99%) | 92% (87-97%) |

**Supplementary Figure 6. Deriving optimal heuristic cut-off thresholds to optimise for sensitivity in Method 2.** A. Method 2 applied to the training, testing and full dataset: Read and percentage cut-off maximising Sensitivity in the training set (rather than for Youden’s index). B. Method 2 applied to the entire dataset: Read and percentage cut-off maximising for Sensitivity (rather than for Youden's Index).

A.

| **Percentage cut-off** | **Read cut-off** | **Sensitivity**  **(95% CI)** | **Specificity**  **(95% CI)** | **Youden's Index** |
| --- | --- | --- | --- | --- |
| 0.4 | 50 | 100% (98-100%) | 100% (97-100%) | 100% |
| 0.4 | 1000 | 100% (98-100%) | 100% (97-100%) | 100% |
| 0.4 to 1.2 | 50 to 100 | 100% (98-100%) | 100% (97-100%) | 100% |
| 1.2 | 50 | 100% (98-100%) | 100% (97-100%) | 100% |
| 1.2 | 1000 | 100% (98-100%) | 100% (97-100%) | 100% |

B.

| **Dataset** | **Percentage cut-off** | **Read cut-off** | **Sensitivity**  **(95% CI)** | **Specificity**  **(95% CI)** | **Youden's Index** |
| --- | --- | --- | --- | --- | --- |
| Training | 0.4 | 50 | 100% (98-100%) | 100% (97-100%) | 100% |
| Testing | 0.4 | 50 | 100% (96-100%) | 100% (95-100%) | 100% |
| Full set | 0.4 | 50 | 100% (98-100%) | 100% (98-100%) | 100% |

C.

| **Percentage cut-off** | **Read cut-off** | **Sensitivity (95% CI)** | **Specificity (95% CI)** | **Youden's Index** |
| --- | --- | --- | --- | --- |
| >0.1-0.3 | 50 | 100% (98-100%) | 99.5% (97-100%) | 99.5% |
| >0.1-0.3 | 1000 | 100% (98-100%) | 99.5% (97-100%) | 99.5% |
| 0.4 | 50 | 100% (98-100%) | 100% (98-100%) | 100% |
| 0.4 | 1000 | 100% (98-100%) | 100% (98-100%) | 100% |

**Supplementary Figure 7. Deriving optimal heuristic cut-off thresholds in Method 2 after adjusting for plausible infections.** A. Method 2 applied to the training dataset after adjusting for plausible infections: Read and percentage cut-off maximising for Youden's Index. B. Method 2 applied to the training, testing and full dataset after adjusting for plausible infections: Read and percentage cut-off maximising for Youden's Index. C. Method 2 applied to the entire dataset after adjusting for plausible infections: Read and percentage cut-off with lower threshold cut-offs.

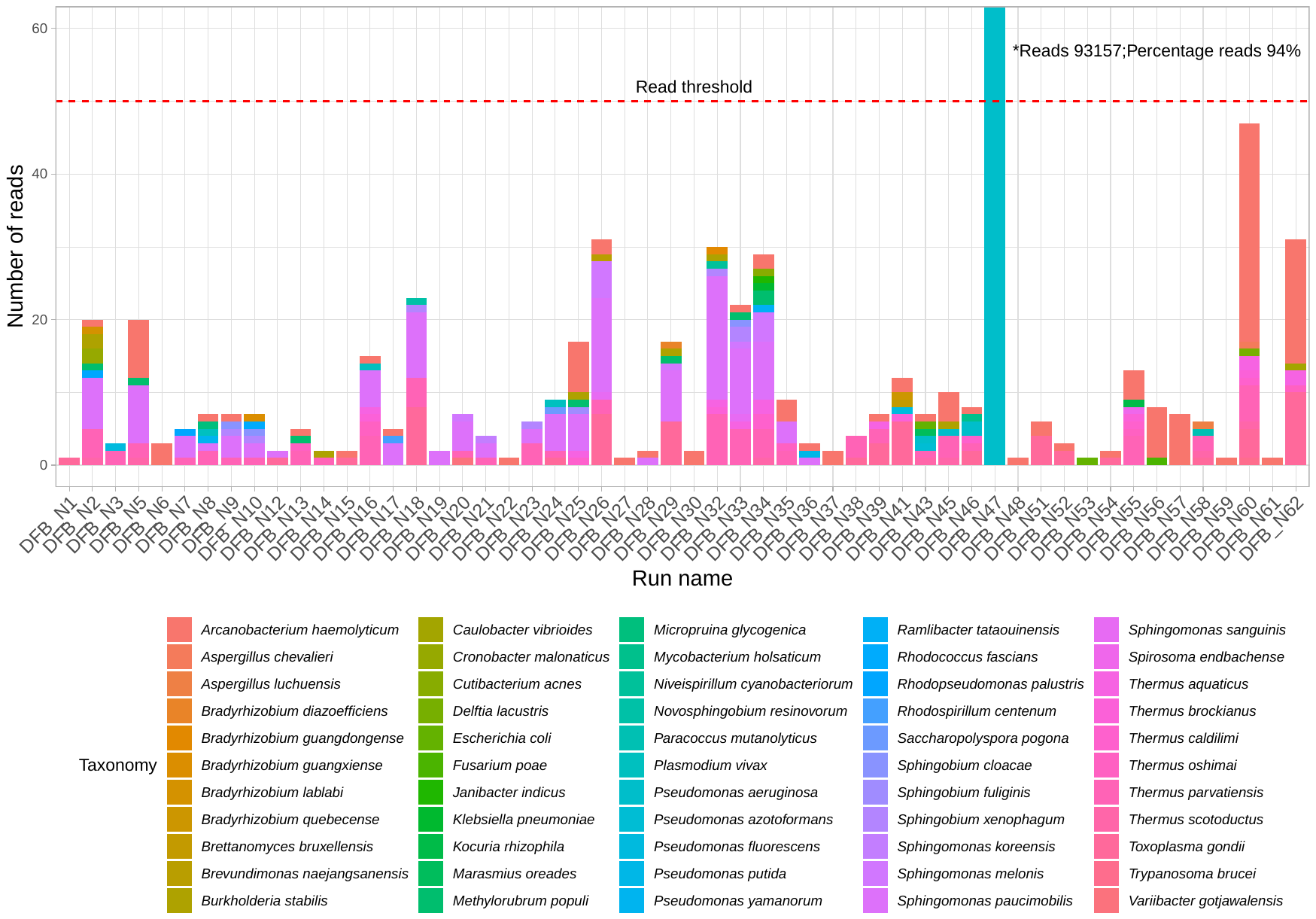

Supplementary Figure 8. Pre-filtered culture negative clinical samples (filter thresholds not applied). Minor cross-contamination which fell well below the thresholds for filtering (i.e., >50 reads) was evident particularly in samples that contained *Arcanobacterium haemolyticum*, which was the organisms used as a positive control in this study. DFB_N47 was shown to be a true infection (*Pseudomonas aeruginosa*) by sequencing and repeat culture.

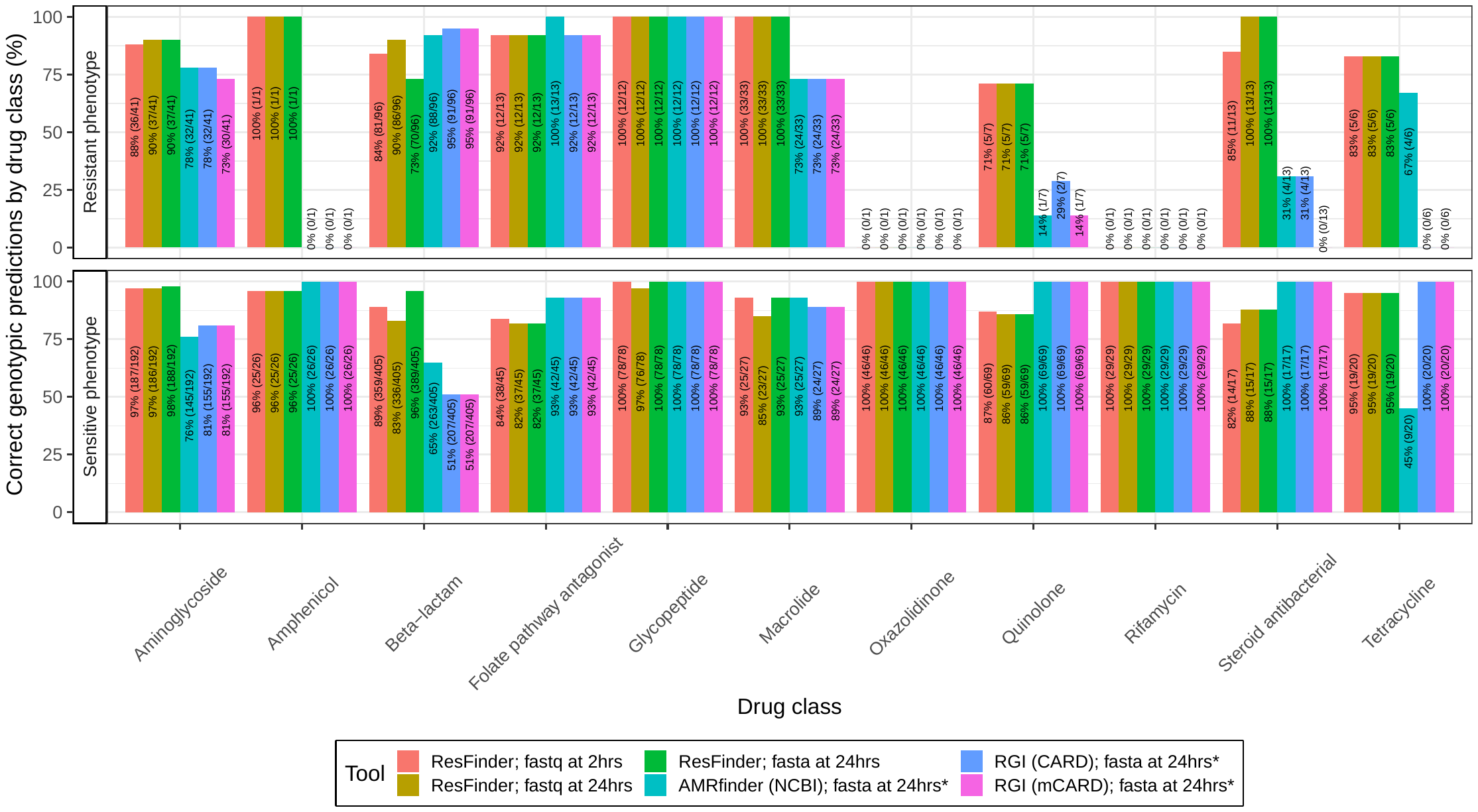
Supplementary Figure 9. Genotypic performance by drug class. Three commonly used AMR prediction software tools were compared. Any AMR predictions found at the drug level was converted to the class level to enable benchmarking of tools.

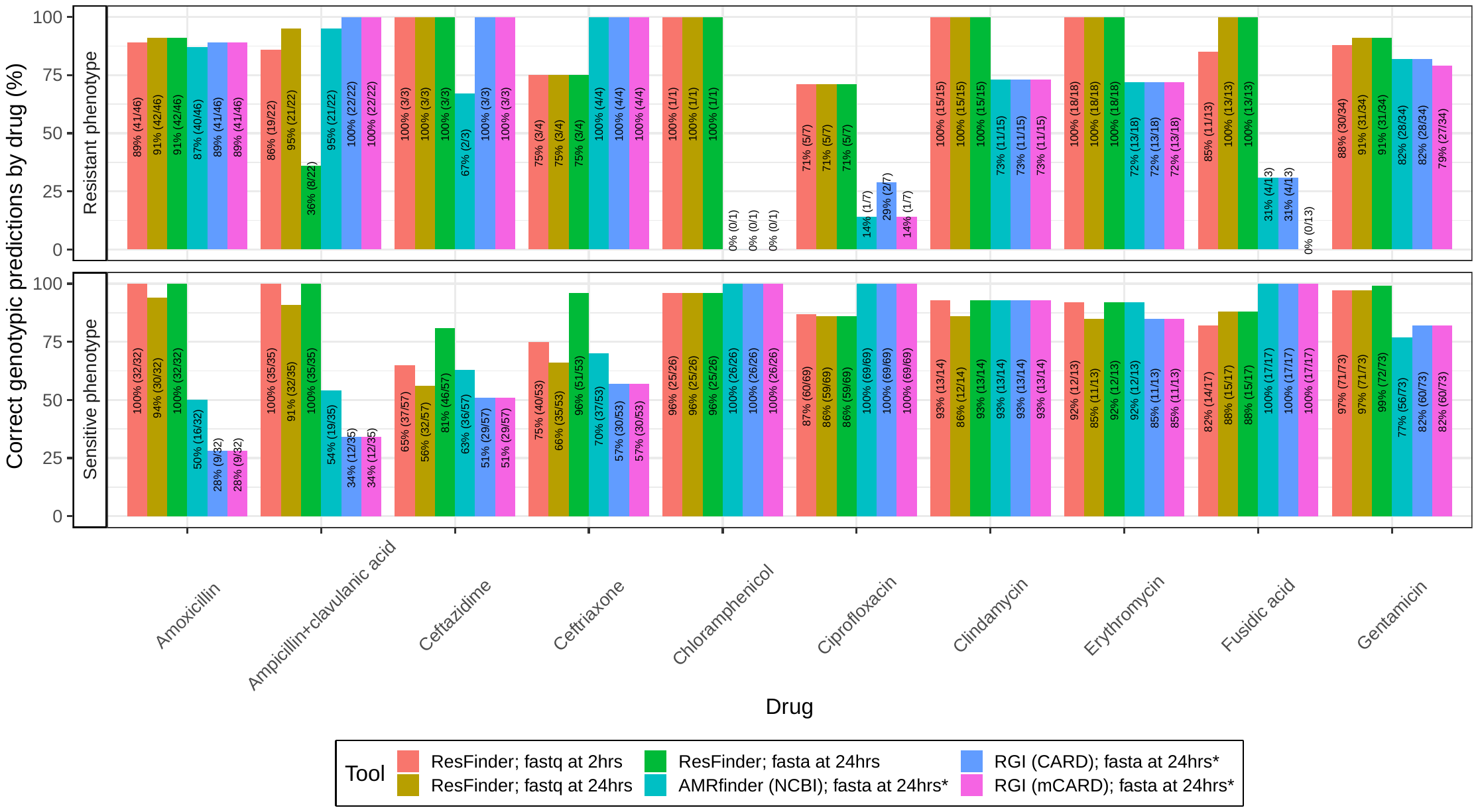
Supplementary Figure 10. Genotypic performance by specific drug (1/2). Three commonly used AMR prediction software tools were compared.

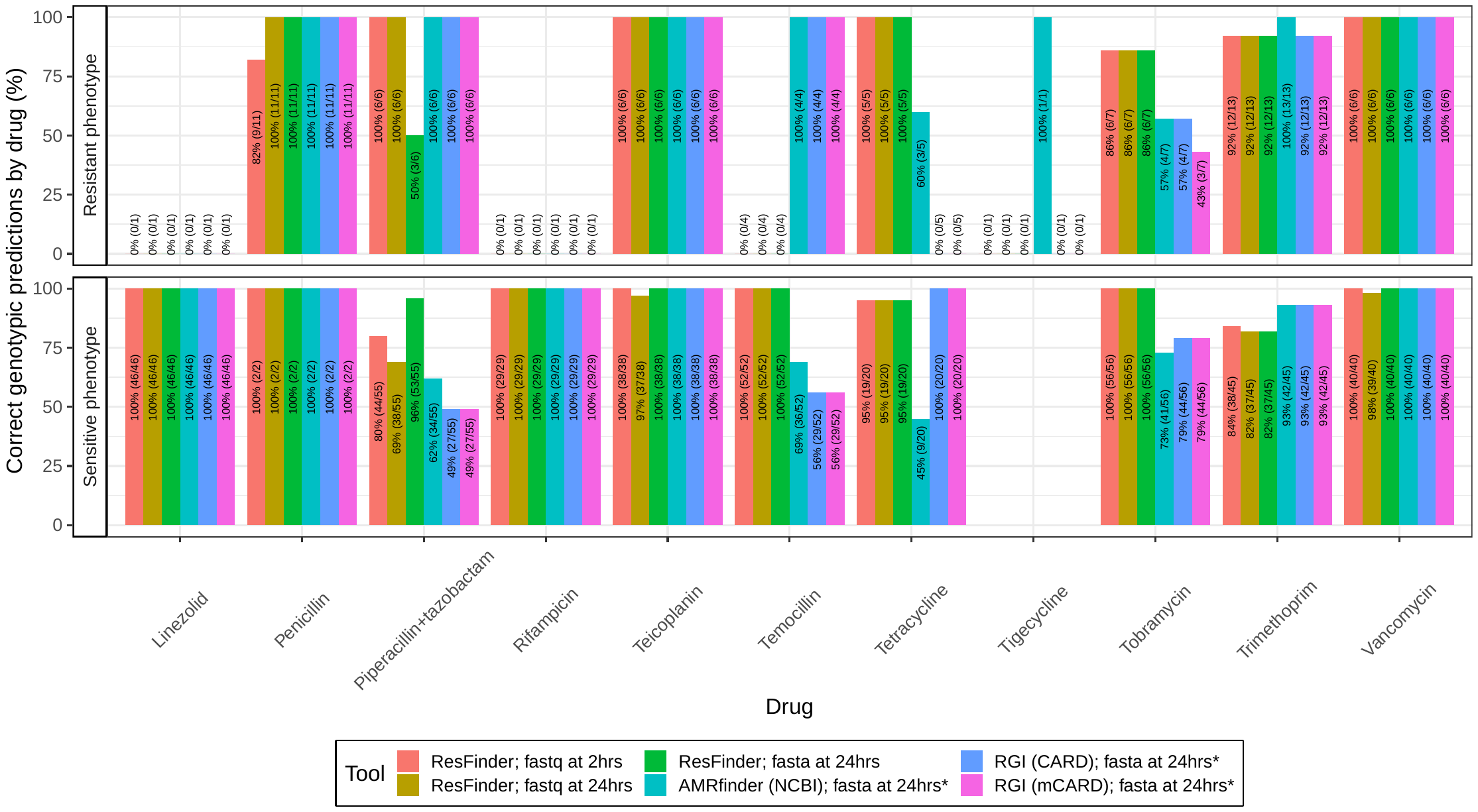
Supplementary Figure 11. Genotypic performance by specific drug (2/2). Three commonly used AMR prediction software tools were compared.

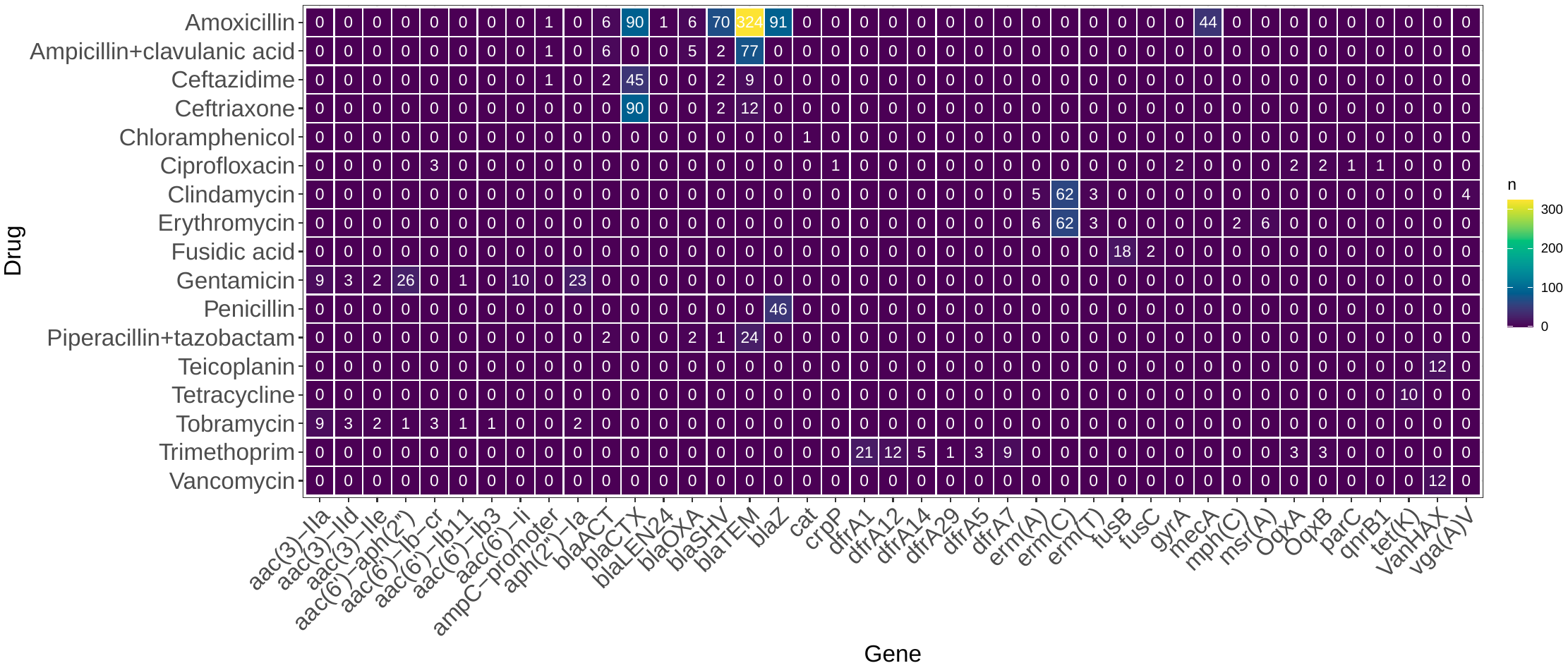

Supplementary Figure 12. Genes implicated in true genotypic calls (ResFinder with fastq reads at 24hr). For conciseness, variants of the genes *blaSHV, blaTEM, blaCTX, blaOXA, blaACT*, and *cat* have been consolidated (detailed in Tables S14, S15).

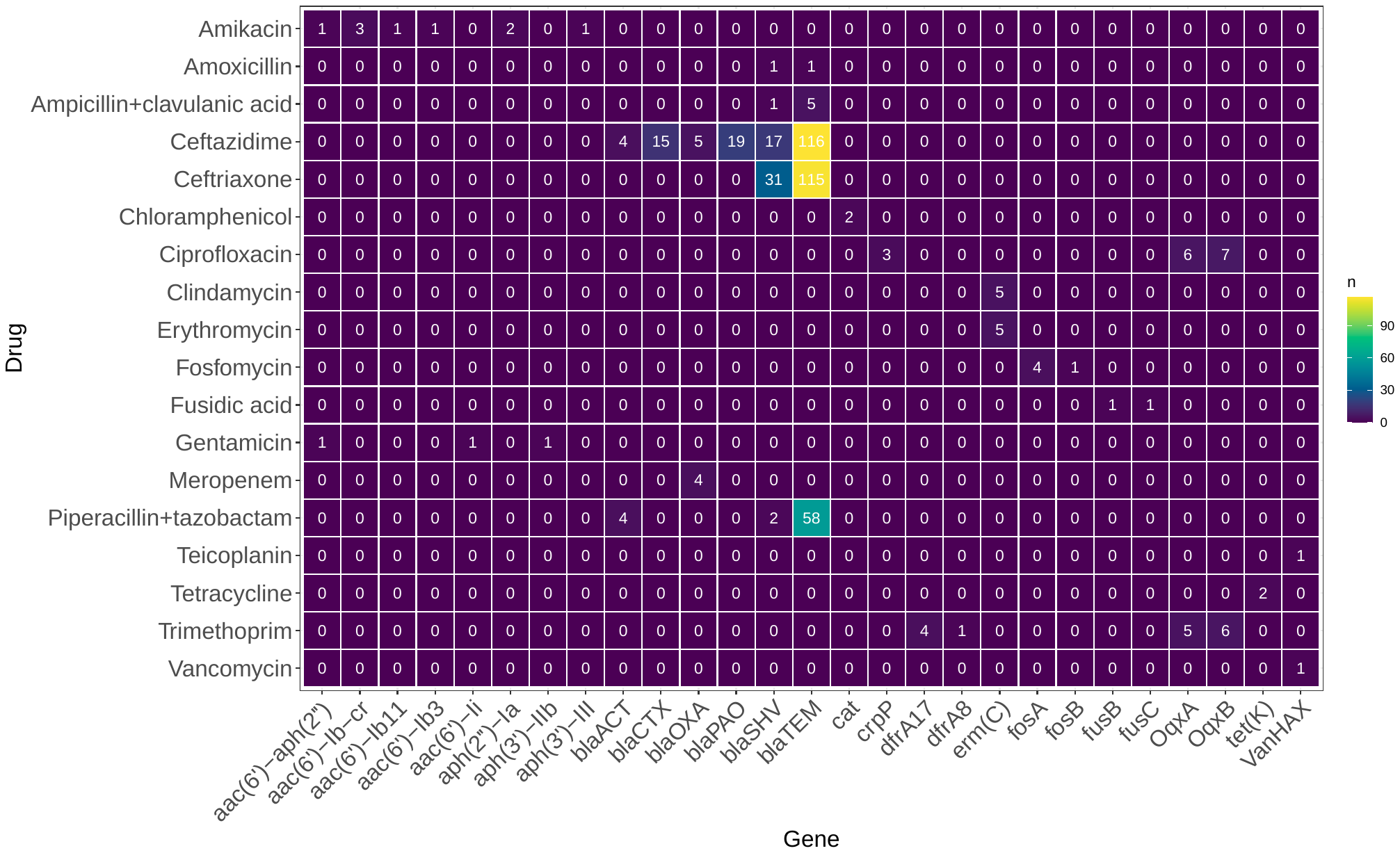

**Supplementary Figure 13. Genes implicated in major error genotypic calls (ResFinder with fastq reads at 24hr).**	For conciseness, variants of the genes *blaSHV, blaTEM, blaCTX, blaOXA, blaACT*, and *cat* have been consolidated (detailed in Tables S14, S15).

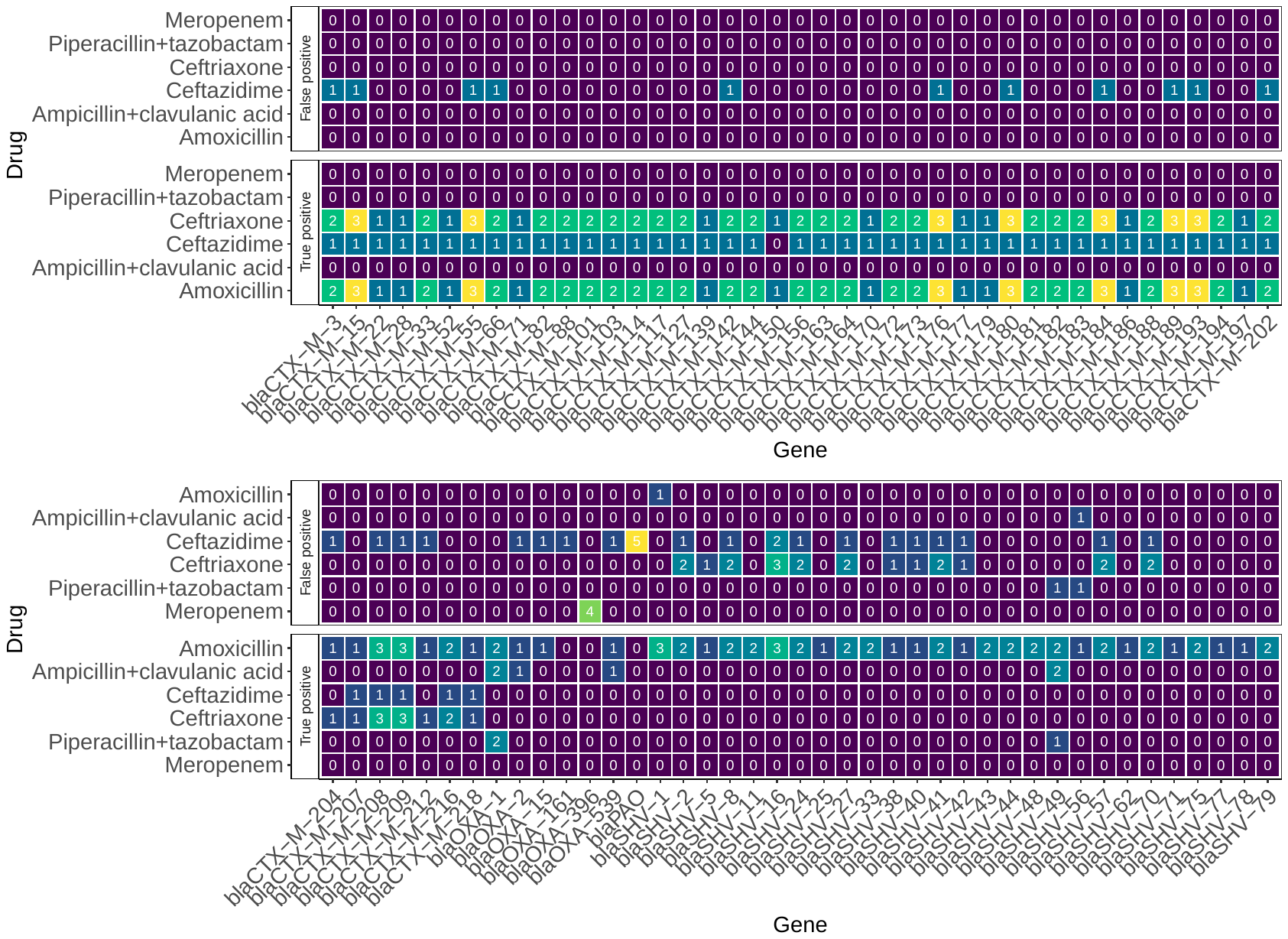

**Supplementary Figure 14. Specific *bla* genes implicated in false and true positive genotypic calls (1/2).**

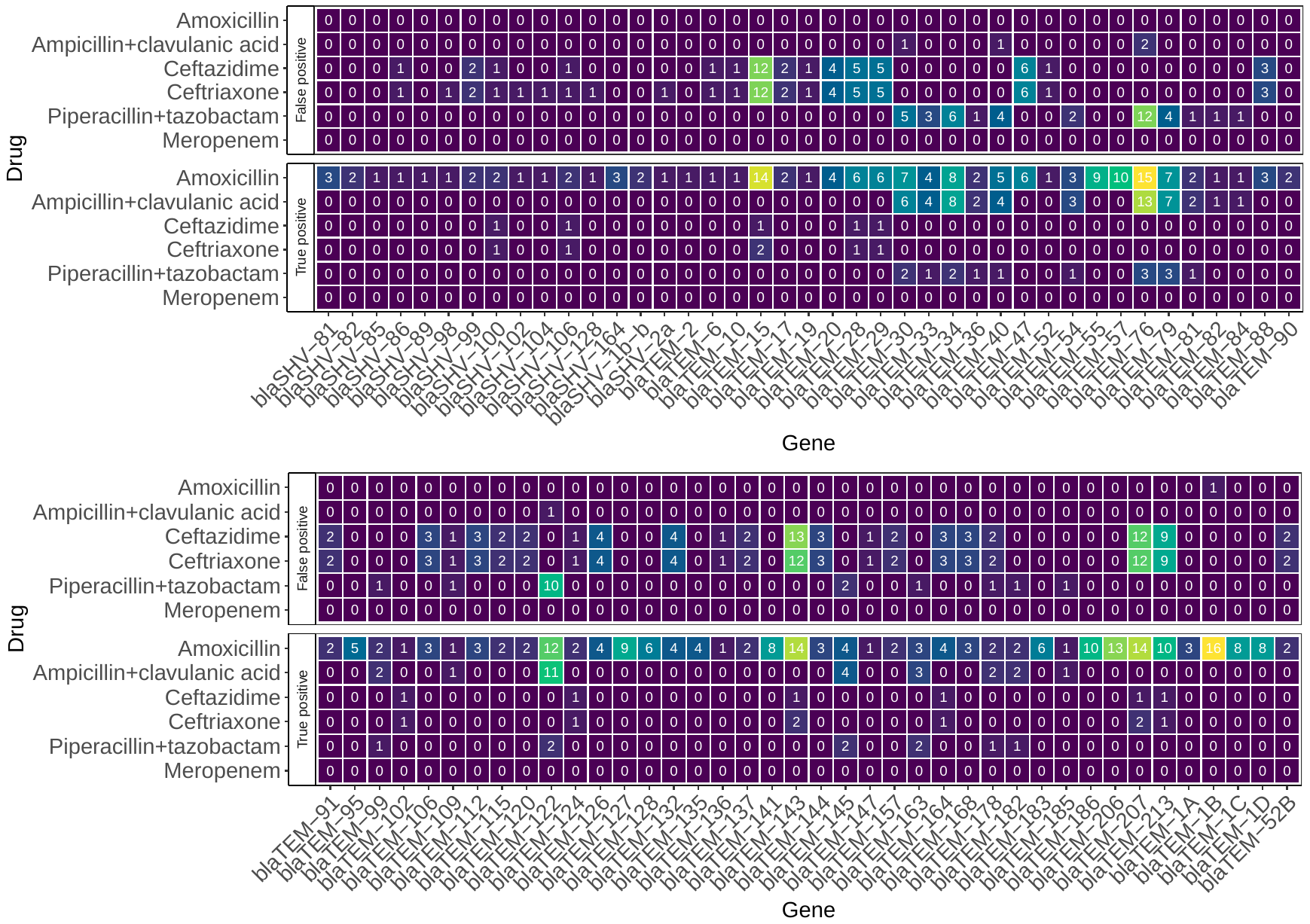
**Supplementary Figure 15. Specific *bla* genes implicated in false and true positive genotypic calls (2/2).**

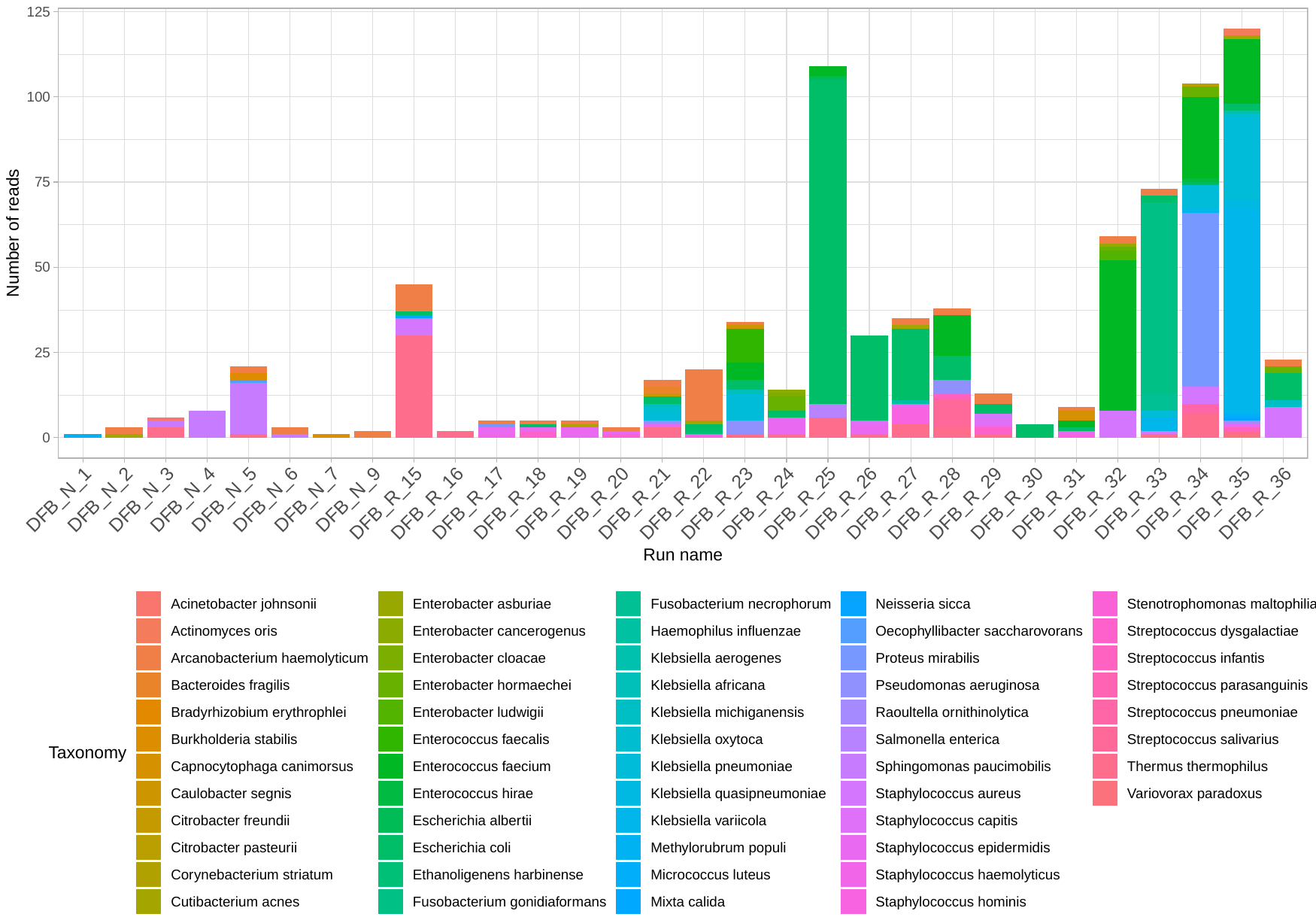

**Supplementary Figure 16. Pre-filtered negative control with no spike (no filter thresholds applied).** “DFB_N” refers to culture-negative samples, whereas “DFB_R” refers to culture-positive samples. Runs are not displayed if there were no reads in the negative control.

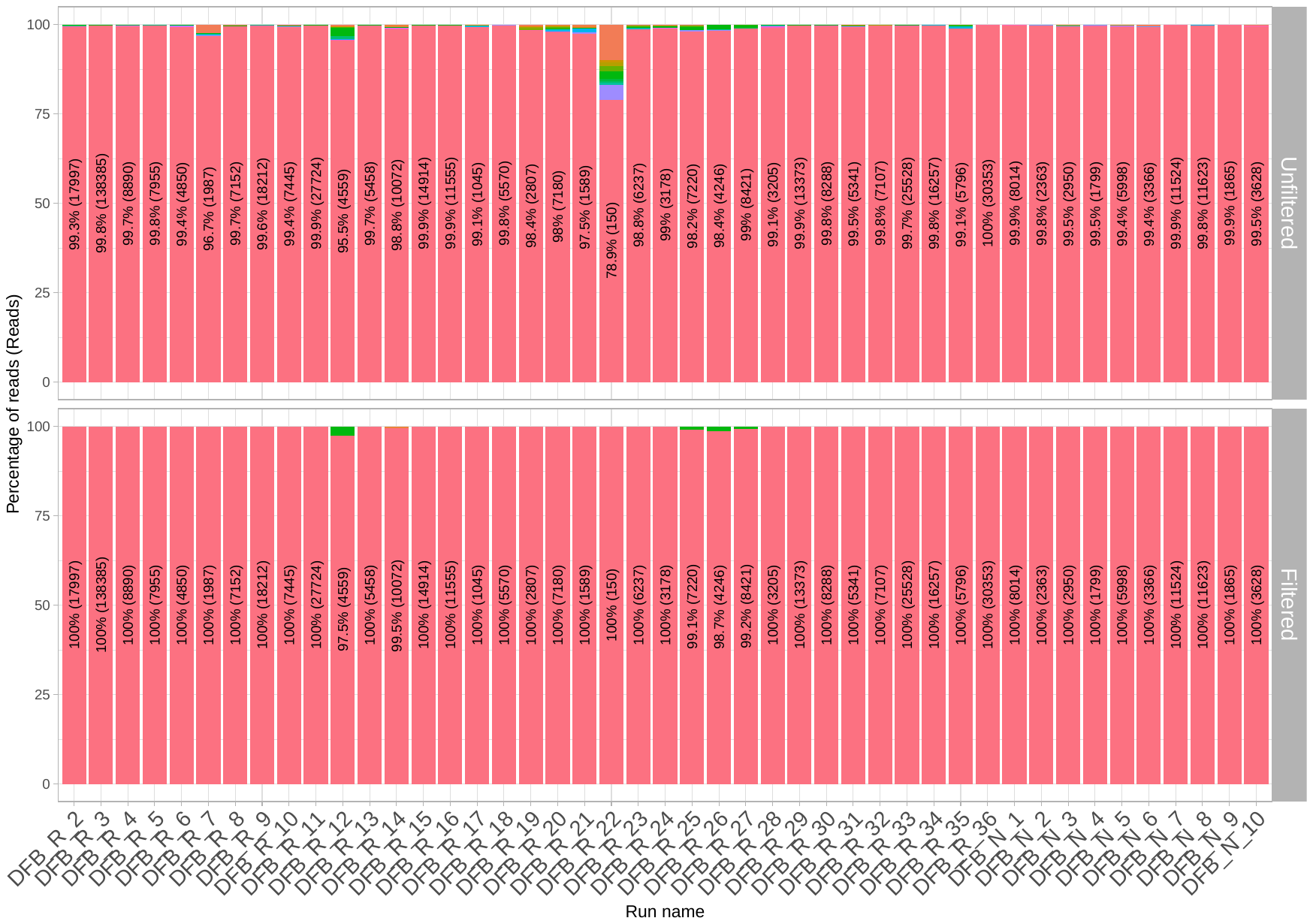

**Supplementary Figure 17. Unfiltered and filtered spiked negative control.** Red bars correspond to *Thermus thermophilus.* The percentage of internal control is shown with absolute read numbers in brackets. The negative control from batch 1 (DFB_R_1) has been removed in the figures, as lambda DNA was added as the spike instead of T. thermophilus in this initial run, before the possibility of mis-identification of lambda DNA as *E. coli* was identified.

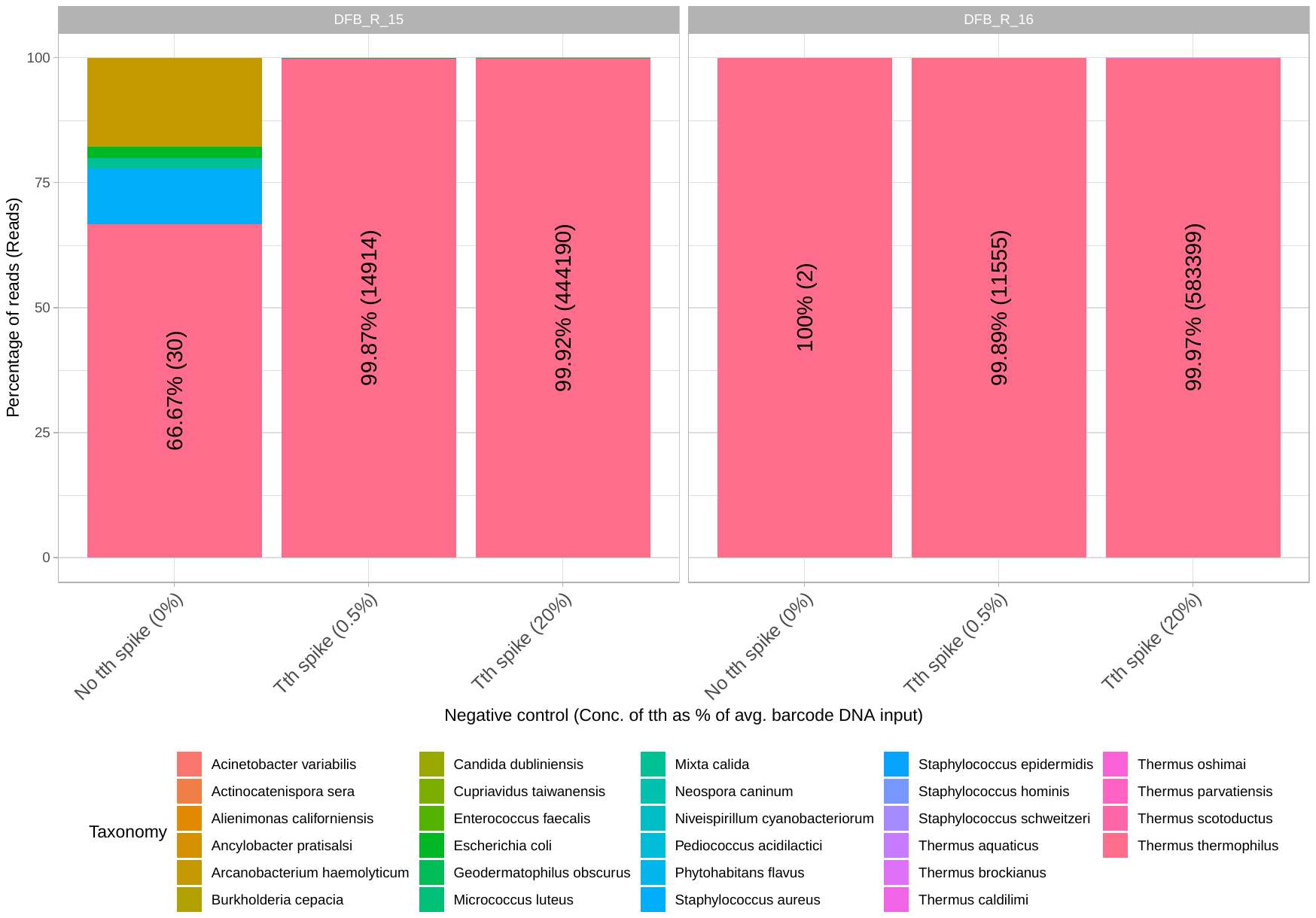

Supplementary Figure 18. Increasing levels of spiked *Thermus thermophilus (Tth)* concentration improves contamination control. Percentage of spike shown with absolute read numbers shown in brackets.

**Supplementary Figure 19. Unfiltered and filtered positive controls.** The percentage of positive control is shown with absolute read numbers in brackets.

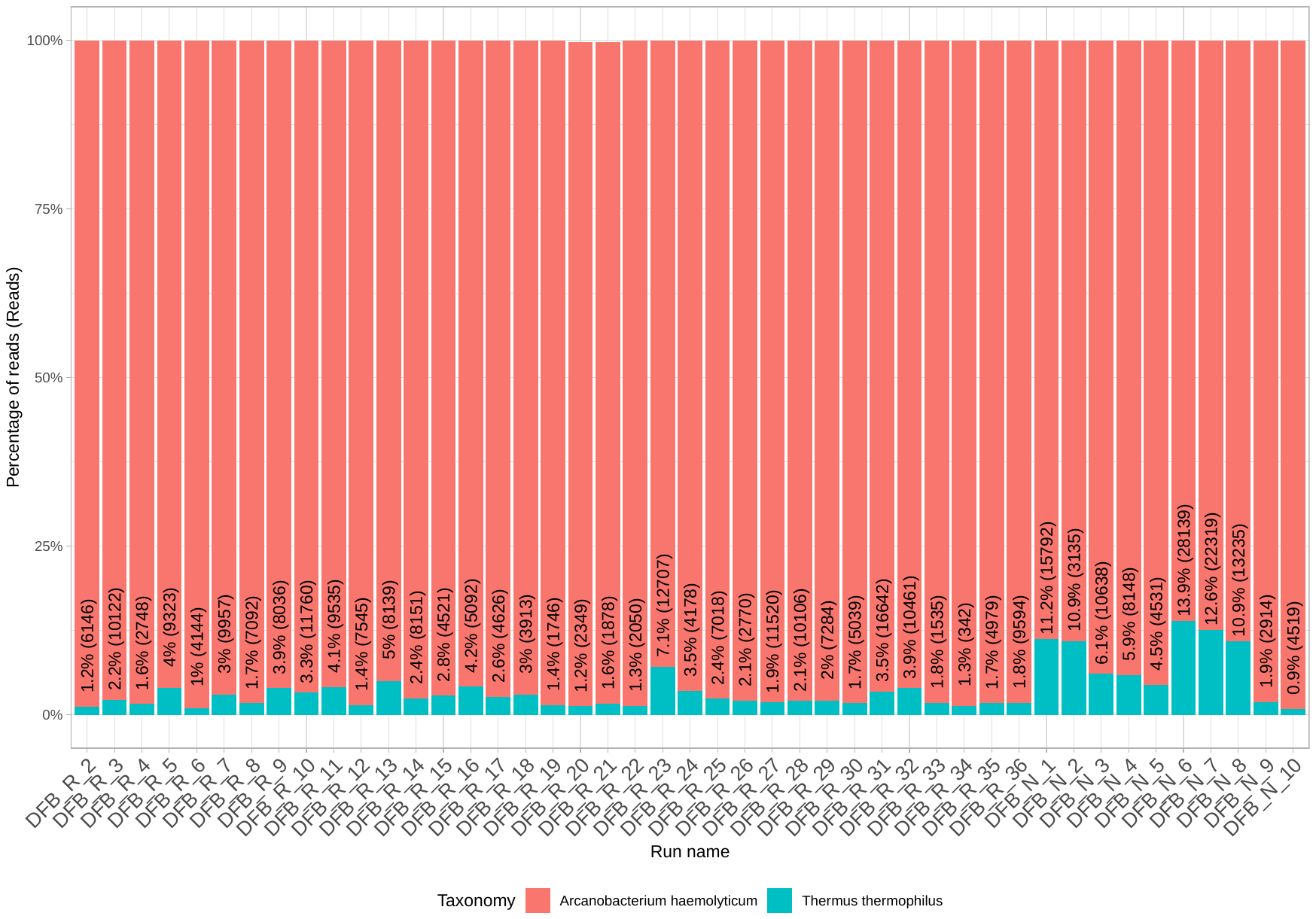

Supplementary Figure 20. Unfiltered and filtered positive controls. The percentage of positive control is shown with absolute read numbers in brackets. The internal control *T. thermophilus* was spiked at 2% of total DNA after DNA extraction, except where the total sample DNA amount was less than 50ng or below the limit of detection (i.e., 2ng/mL), when a fixed amount of 0.5ng DNA was added as the internal control.

Supplementary Figure 21. Internal spike control shown in culture-positive clinical samples. The percentage of internal control is shown with absolute read numbers in brackets. The internal control was spiked at 2% of total DNA after DNA extraction using the Qubit 2.0 fluorometer (broad range kit; Thermo Fisher Scientific, USA). In cases where the total positive control DNA amount was less than 25ng, a fixed amount of 0.5ng *T. thermophilus* DNA was added as the internal control, as lower amounts become difficult to reliably quantify. Therefore, in these select samples *T. thermophilus* would make up >2% of total reads.

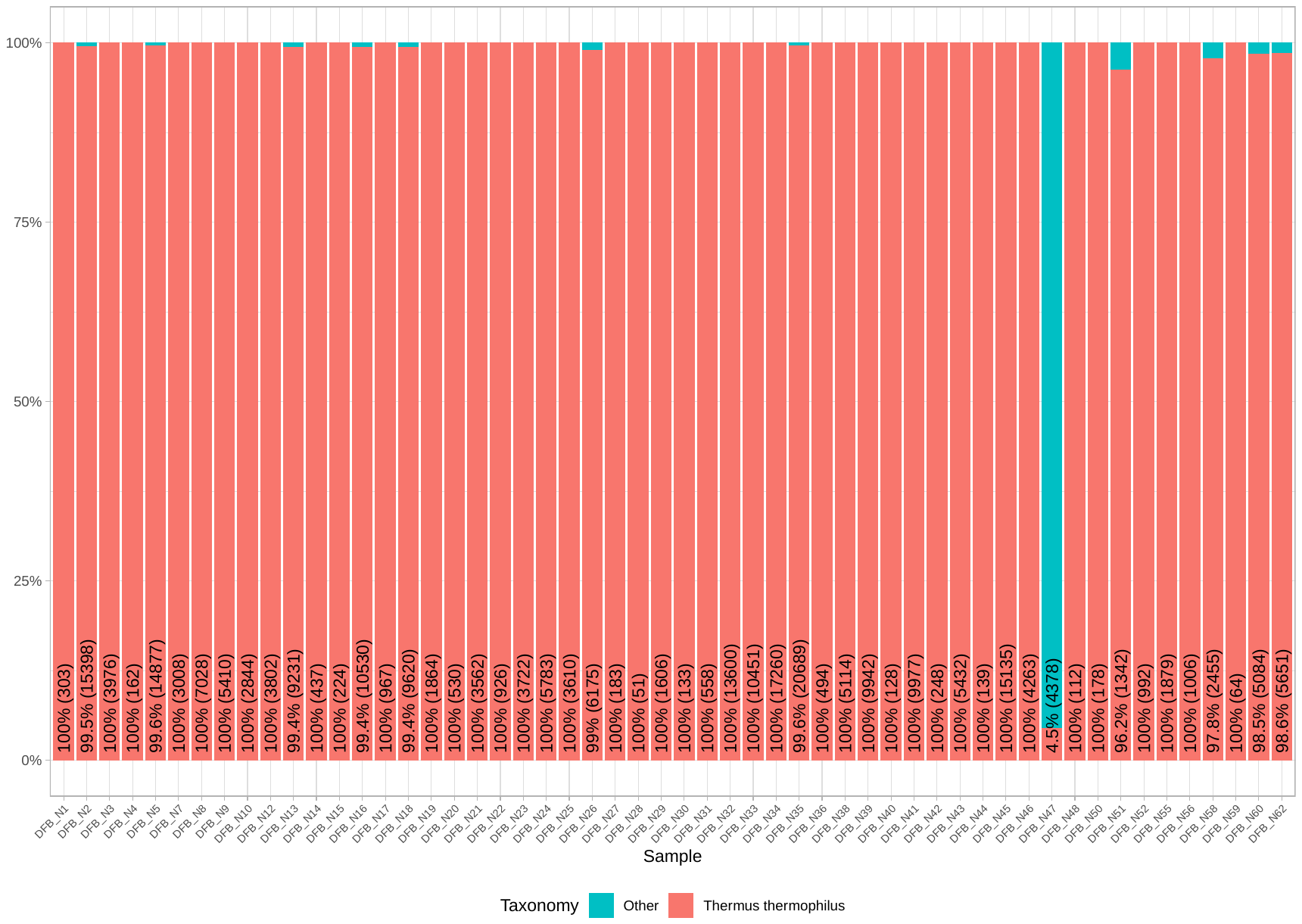

Supplementary Figure 22. Internal spike control shown in culture-negative clinical samples. The percentage of internal control is shown with absolute read numbers in brackets. Clinical negative cultures contained generally 100% of the internal control *T. thermophilus*, except for sample N47 which found to contain *Pseudomonas aeruginosa* by sequencing and confirmed by repeat phenotypic testing.

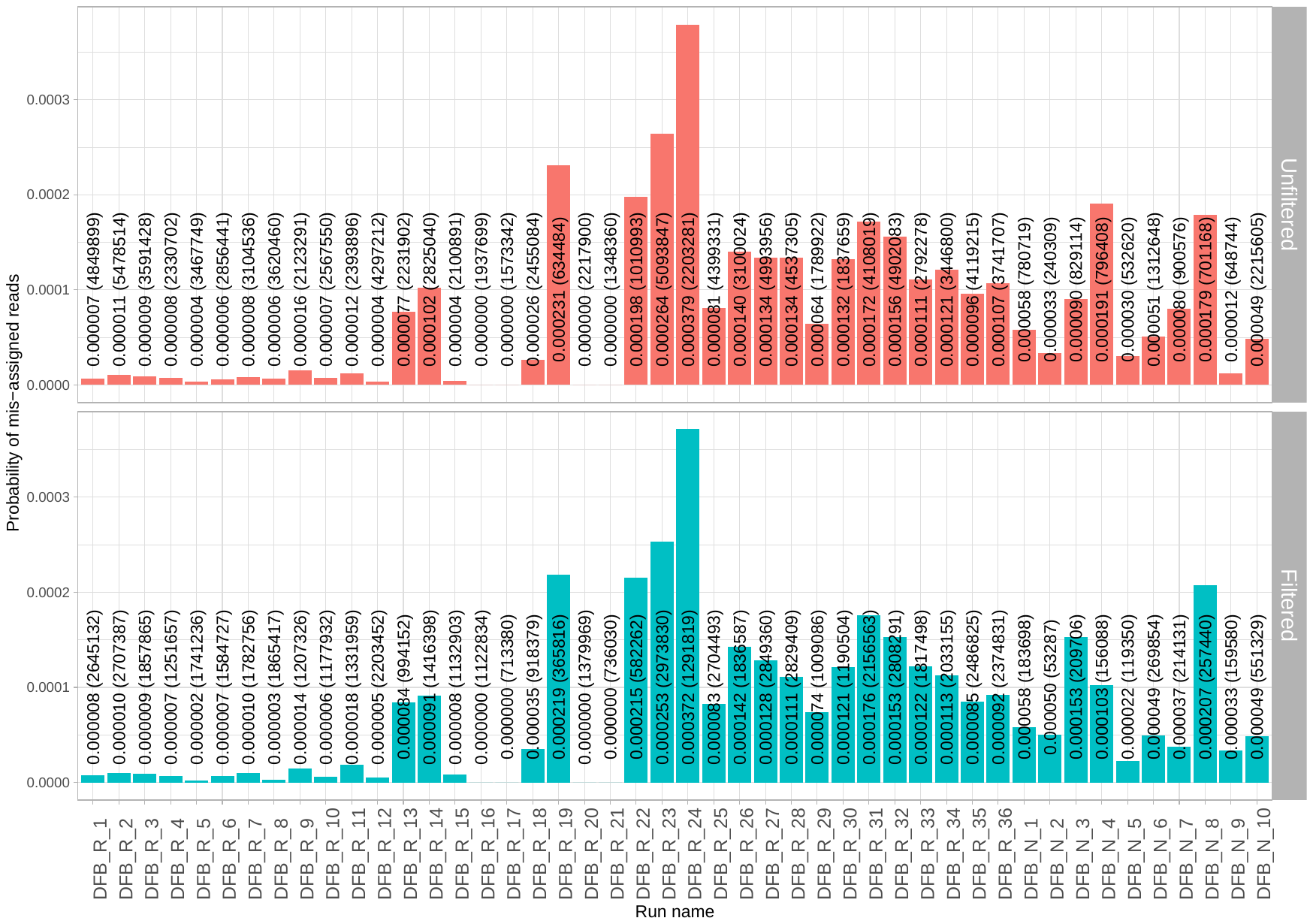

Supplementary Figure 23. Probability of reads mis-assigned to other barcodes per run. The mean probability of mis-assigned reads is shown with the absolute number of reads in brackets.

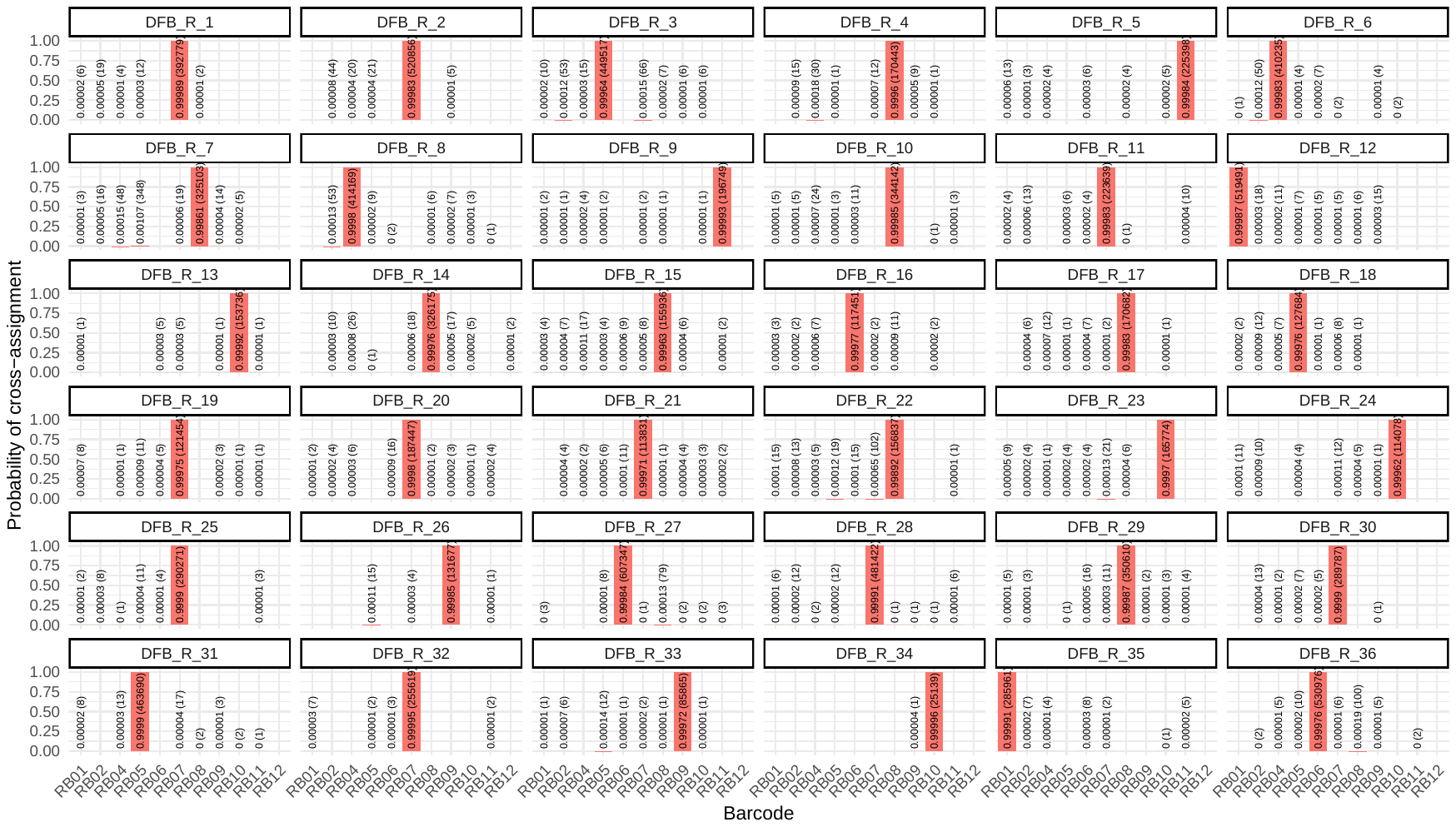
**Supplementary Figure 24. Probability of cross-assignment in culture positive samples (estimated using *Arcanobacterium haemolyticum*).** The largest red bar in every run corresponds to the correct positive control sample. The mean probability of cross-assigned reads is shown with the absolute number of reads in brackets.

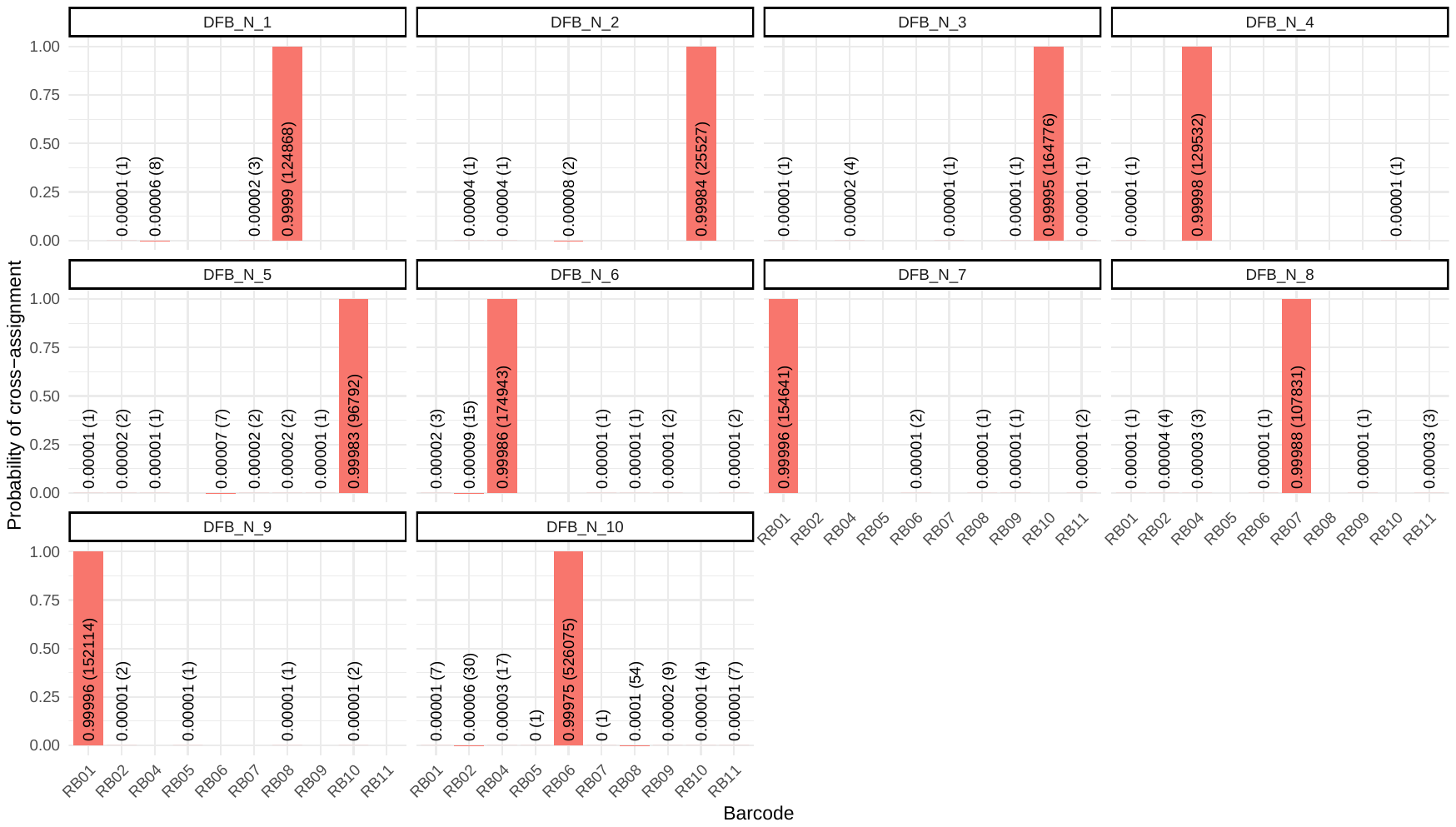

**Supplementary Figure 25. Probability of cross-assignment in culture negative samples (estimated using *Arcanobacterium haemolyticum*).** The largest red bar in every run corresponds to the correct positive control sample. The mean probability of cross-assigned reads is shown with the absolute number of reads in brackets.

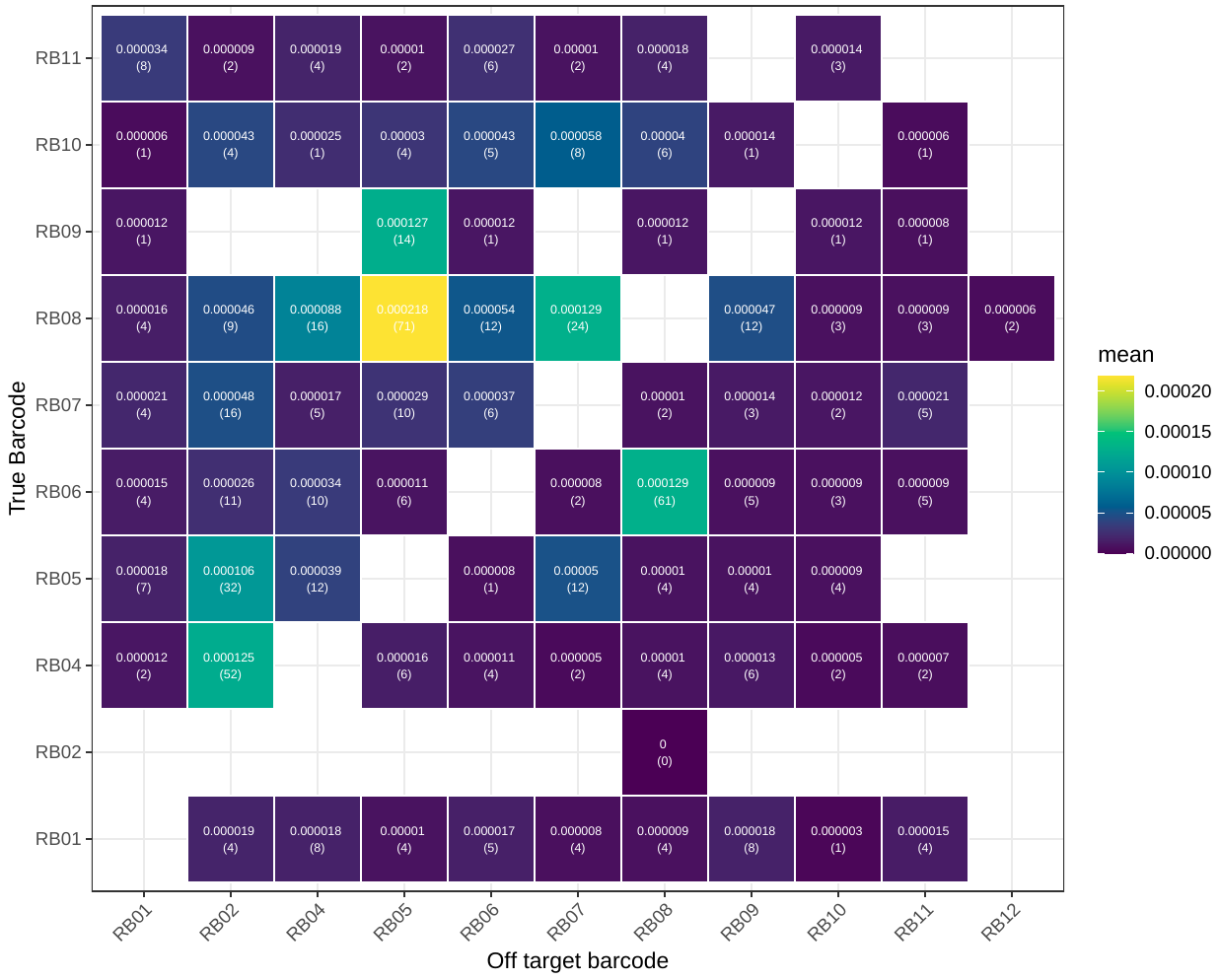

Supplementary Figure 26. Cross-assigned *Arcanobacterium haemolyticum* reads shown across off-target barcodes. The mean probability of cross-assigned reads is shown with the absolute number of reads in brackets.
