## Supplementary Methods and Results for "Rapid clinical diagnosis and treatment of common, undetected, and uncultivable bloodstream infections using metagenomic sequencing from routine blood cultures with Oxford Nanopore"

#### Sample Collection and Processing

At the point of extraction, colony forming units (CFUs) were determined using chocolate, blood and chromagenic agar plates using standard conditions. Following incubation, colonies were counted manually, and final CFU counts were recorded as the median across all three agar types, ensuring accuracy and minimising plate-specific bias.

#### DNA extraction from blood culture

As a positive control for each batch, we used a single 5ul loop of *Arcanobacterium haemolyticum* grown from blood agar plate cultures. Extracted DNA from clinical samples were normalised prior to library preparation, and DNA from positive controls were normalised to one-fourth of this concentration to ensure greater sequencing coverage depth of clinical samples. A spiked internal control of *Thermus thermophilus* DNA at a concentration of 2% of the normalised DNA concentration (with an absolute minimum amount of 0.5ng) was added to every clinical sample, the positive and one negative control, leaving the other negative control completely free from any DNA input.

#### Bioinformatic analysis

##### Species prediction

A full standard database was built from archaeal, bacterial, plasmid, viral and human reference genomes downloaded from NCBI RefSeq on the February 13^th^, 2022 (Figure S2).

Heuristic cut-off threshold models (Method 1 and 2) and random forest model (Method 3) were derived to identify the presence of true infection. We used 60% of the culture bottles processed as training/derivation data and the remaining 40% as the testing set (selected at random using the sample_frac() function in RStudio (V 2023.03.0+386)). The same split was used throughout all methods tested to ensure comparable results. For Methods 1 and 2, optimal thresholds for determining if samples contained low-level contamination or true infection from sequencing were determined by numerical optimization in the training set, choosing thresholds that maximised the Youden's Index (Sensitivity + specificity - 1). In Method 3 we trained a random forest classifier.

The first method removed plasmid reads prior to classification using a plasmid only Kraken2

database that was built (sequences downloaded from NCBI on the March 16^th^, 2023), as plasmids, due to horizontal gene transfer, may be classified into multiple species erroneously. To derive cut-off thresholds, the following filters were used for Method 1: read counts and percentage cut-off, that is, the total number of reads classified as a specific species, and the percentage of all bacterial reads classified as a specific species.

The second method did not remove plasmid reads, but instead used Minimap2 (v2.24-r1122; using the map-ont function) to map sequencing reads to each species identified by Bracken to a species-specific reference genome (downloaded randomly from NCBI) after minimal filtering (<0.1% percentage of reads) that excluded only the least plausible species before mapping. Samtools (v 1.17) was used to determine 3 additional metrics that were used for filtering: coverage breadth, depth of coverage, and percentage of reads mapped from extracted species reads. Heuristic thresholds were derived for all 5 filtering metrics from the training set and the applied to the test set. The latter three, coverage breadth, depth of coverage and percentage of reads were set to the lowest values for a true species identified in the training set and the remaining thresholds set optimising the Youden index as above. Following an initial evaluation, thresholds were retrained using a "corrected" training dataset, correcting for plausible false-positive and false-negative errors that were identified.

The third method, used the same 5 metrics as inputs, but applied a machine learning approach using a random forest model to classify Bracken and mapping results. The randomForest package in R developed by A. Liaw et al was used.^1^ Hyperparameter selection was made using a repeated cross-validation approach within the training data with 5 folds and 40 repeats. A random forest model was trained using the training set, and its variable importance was assessed. The model was evaluated on the testing set.

##### Assessing false negative Method 2 results for plausibility (i.e., potential inaccurate sequencing results or inaccurate MALDI-TOF results)

To interpret MALDI-TOF results on cultured isolates, probability scores are used: a value of >2.3 indicates “highly probable species identification”, a score >2 and <2.3 indicates “secure genus identification, probable species identification”, a score >1.7 and <2 indicates “probable genus identification”, and a score <1.7 indicates “unreliable identification”. ^2^

##### Assessing false positive Method 2 results for plausibility (i.e., potential inaccurate sequencing results or inaccurate MALDI-TOF results)

False positive results, that is, additional species detected from sequencing not found on routine culture, were assessed for bioinformatic plausibility. Firstly, 1000 reads were extracted from additional species detected and queried using NCBI BLAST (v2.13.0+) with the ref_prok_rep database (downloaded March 12^th^, 2023) to see if the same species was found. Hits were filtered using percent identity>=95, coverage >=80, E-value <1e-50 and the -max_target_seqs option set to 1. Secondly, for each additional species identified by Bracken, extracted reads were mapped using Minimap2 (v2.24-r1122; using the map-ont function) to a randomly selected refseq genome of the same species (downloaded from NCBI April 29^th^, 2023). Samtools (v1.17) was used to calculate genome coverage breadth and depth. To investigate the extent to which bioinformatic misclassification might arise between genomically similar species, the Average Nucleotide Index (ANI) was calculated for additional bioinformatic species found against the MALDI result using ANI.pl (downloaded reference genomes from NCBI April 29^th^, 2023).^3^ Additional microbiology data was captured from other blood culture samples, if any were available, within 5 days of the initial collected blood sample to look for evidence of the additional species in these samples. Laboratory requests were also reviewed for relevance to additional infections found by sequencing. Provision for this is included in the ethical approvals cited above.

#### Statistical analysis

The unadjusted performance of metagenomic sequencing was assessed by comparing the species identified from sequencing data with the species from pure culture isolates grown from blood culture samples [species from Bruker microflex MALDI-TOF MS (Bruker, USA)], while the adjusted performance additionally considered plausible infections identified bioinformatically. Sensitivity was calculated taking each species identified from each culture-positive sample as a separate data point contributing to the numerator and denominator; therefore culture-positive samples with a single species contributed once, and culture-positive samples with two or more species contributed two or more times. Specificity was determined both by considering the proportion of samples deemed negative by blood culture that were identified as negative by metagenomic sequencing, and in culture-positive samples. In the latter samples false-positive results are still possible where an additional species not found by culture was reported. In culture positive results, specificity was defined on a per sample basis, where any sample with >1 additional species found from sequencing was considered to contribute as 1 to the false negative count.

#### Antimicrobial resistance prediction

Results that were determined to be "intermediate" was excluded from the analysis. To ensure higher quality predictions and to keep plasmid sequences, sequencing data was pre-processed by filtering out human and *Thermus thermophilus* reads prior to resistance prediction using the Kraken Software Suite, rather than extracting only identified species reads.^4^ Genotypic resistance prediction was performed with fasta assemblies [Flye v. 2.9-b1768 with '-meta' and '-nano-raw' ] and raw reads when possible using ResFinder 4.0 (ResFinder & PointFinder db downloaded July 3^rd^, 2022), Resistance Gene Identifier (CARD downloaded August 18^th^, 2022), and NCBI AMRFinderPlus 3.10.40 (db downloaded July 30^th^, 2022). Additionally, a curated mapping database (that maps genes found to genotypic resistance) from ResPipe, an open-source software pipeline, was also applied to RGI (CARD) hits to determine genotypic resistance predictions which is referred to as ResPipe (modified CARD, mCARD).^5^

RGI(CARD) was configured to excluded hits associated with the "antibiotic efflux" mechanism and the "protein variant model". The former, a nonspecific mechanism influenced by factors such as efflux pump expression levels, and the latter, a context-dependent model susceptible to variability among different bacteria and conditions, were filtered out to focus on more specific and reliable resistance mechanisms. We set stringent thresholds for best identities (>60), best hit bitscore (>33), and percentage length of reference sequence (>99) to ensure significant sequence similarity, high-quality alignment, and comprehensive coverage of the reference sequence, respectively.^6,7^ For AMRFinder, we set a percentage coverage of the reference sequence to >85%, percentage identity to the reference sequence >95% and minimum target length >120bp to ensure that the identified sequences likely represent substantial portions or complete genes, rather than fragments, reducing the risk of false positives from partial gene matches.^6,7^ We also filtered for the "AMR" (Antimicrobial Resistance) subtype, focusing the analysis on elements specifically associated with antimicrobial resistance and excluding irrelevant elements.

Of note, we used the *Staphylococcus aureus* rules across all *Staphylococcus* species to ensure a broader coverage and attempt detection of resistance elements. Monomicrobial and polymicrobial infections were analysed separately. In cases where discordant phenotypic susceptibility results existed in polymicrobial species, a genotypic result of resistant was required to correctly predict this phenotype, counting as a single drug prediction.

#### Barcode contamination

Within multiplex sequencing data, reads assigned to barcodes not included in the experiments were defined as mis-assigned reads, and reads assigned to the incorrect barcode bins in-use were referred to as cross-assigned.^8^ In this study, we assessed both mis-assigned reads by Guppy demultiplexer and cross-assigned reads using the positive control *A. haemolyticum* as a proxy. *A. haemolyticum* was used as this was added only to a single positive control sample per run sequenced and this species was not present in any of the MALDI-TOF results from this dataset.

##### Mis-assigned reads

As there are 12 barcodes available, and 11 are in use with barcode number 3 not in use, $number\left( barcode 3 reads \right)= \left( 1- p\left( t \right) \right)\times\left[ \frac{1}{11} \right]\times z$, where p(t): probability of a read being classified to the correct barcode and z: total number of reads. The term 1/11 reflects that when mis-assigned there are 11 possible barcodes that a read can be mis-assigned to. Therefore,

$p(t)= 1-[ \frac{number\left( barcode 3 reads \right)}{z} \times11 ]$.

Now assuming there are more barcodes not in use (a1, a2, …). The equation then becomes:

$$p(t)= 1-[ \frac{number\left( bar. a1 reads \right)}{z} + \frac{number\left( bar. a2 reads \right)}{z}+ ...]\times\frac{(number of total barcodes in use-1)}{(number of barcodes not in use)}.$$

##### Cross-assigned reads

The probability of reads cross-assigned was calculated by the number of *Arcanobacterium haemolyticum* reads not in the positive control barcode, but assigned to a barcode in use, divided by the total *Arcanobacterium haemolyticum* reads present in a run assigned to any barcode in use.

### Supplementary results

#### Results: Species identification

##### False-negative results (Method 2)

In samples 22, 23, 46 and 49, MALDI-TOF predicted *Corynebacterium lipophiloflavum, Staphylococcus epidermidis, Klebsiella* *pneumoniae* and *Klebsiella pneumoniae (MALDI scores of 2.08, 2.07, 2.35, and 2.35 respectively)* while sequencing results predicted *Corynebacterium sanguinis*, *Staphylococcus hominis, Klebsiella quasipneumoniae* and *Klebsiella* *quasipneumoniae* respectively. Samples were repeat tested grown from saved stock cultures and yielded the same MALDI-TOF results as described. In all four samples, there was higher coverage breadth of the sequencing reference genome species than the MALDI-TOF reference genome species (93% vs 92.7%, 95% vs 63%, 62% vs 59% and 68% vs 62%, Table S3). Furthermore, almost all blast hits were classified to the species identified by sequencing (99.2%, 97%, 96% and 96% respectively) rather than the MALDI-TOF species. Prior studies have shown that *Corynebacterium* *sanguinis* and *Corynebacterium lipophiloflavum,* and *Staphylococcus epidermidis* and *Staphylococcus hominis* are genotypically similar and are often misidentified by MALDI-TOF.^9–11^ Furthermore, the MALDI-TOF probability scores for these two organisms were 2.08 and 2.07 respectively, suggesting a reliable genus identification, but only probable species identification.

*Klebsiella quasipneumoniae* is a newly defined species, not readily distinguished from *Klebsiella pneumoniae* with current MALDI-TOF identification techniques, which has resulted in its true prevalence being unknown.^12^ This is likely due to a lack of discrimination between spectra patterns being produced by these species. *Klebsiella quasipneumoniae* has been shown to be capable of nosocomial transmission and acquiring and maintaining relevant resistance elements, placing an increased necessity for accurate identification of this organism.^12^ Biomerieux API (Analytical Profile Index) 20E testing was performed on samples 46 and 49 which were both sequenced as *Klebsiella quasipneumoniae,* as well as sample 60 which was sequenced as *Klebsiella pneumoniae* as a control for phenotype. Sample 60 was identified by the API test to have a high probability (i.e., % ID: 82) as being *Klebsiella pneumoniae*, while both samples 46 and 49 were identified with a much lower probability (i.e., % ID: 21) as being identified as *Klebsiella pneumoniae*. This difference in probability score was due to the two samples sequenced as *Klebsiella quasipneumoniae* having a negative LDC result, which tests for decarboxylations of the amino acid lysine by lysine decarboxylase. Furthermore, *Klebsiella quasipneumoniae* is not in the API 20E database, and therefore a much lower probability is expected.

#### Controls

##### Negative control: No spike

Two negative controls were added to each sequencing flow cell, 30 in total. In all negative control samples with no spike added and no bioinformatic filtering, total reads did not exceed 125 (Figure S16).

##### Negative control: *Thermus thermophilus* spike

When *Thermus thermophilus* DNA spikes were added to negative controls (targeting 2% of the normalised DNA concentration [with an absolute minimum amount of 0.5ng]), >96% of the classified reads were from this species, except for one sample (DFB_R_22) which had a low total read number of 150 (Figure S17). After applying filtering thresholds (reads>50, percent reads>0.4%), all samples were classified as having >98% of the spike added, while filtering by reads>150 and percent reads>0.4% resulted in 100% of the spiked-in species in all negative control samples.

##### Negative control: Increasing *Thermus thermophilus* spike levels

In samples 15 and 16, an additional negative control of *Thermus thermophilus* spiked at 20% of the individual sample DNA concentration was added to assess the effect of concentration on contamination. Prior to any filtering, increasing the concentration of *T. thermophilus* DNA from 0% to 0.5% and to 20% improved contamination control and resulted in 66.7%, 99.87% and 99.92% and 100%, 99.89% and 99.97% of reads classified as *T. thermophilus* in both samples respectively (Figure S18).

##### Positive control

Positive controls which consisted of *Arcanobacterium haemolyticum* were present in every run. Unfiltered positive controls had >99.8% of reads classified as *A. haemolyticum*. After applying filtering thresholds (percent>0.4, reads>1000, coverage_breadth>32, reads_mapped>75, mean_depth>0.4), all positive controls had 100% of reads classified to *A. haemolyticum* (Figure S19).

##### Internal spike control present in positive controls

The internal spike of *Thermus thermophilus* DNA was added to all positive controls (2% of the normalised DNA concentration (with an absolute minimum amount of 0.5ng)). The median percent of *T. thermophilus* reads was 2.4% (1.7-4) [0.9-14] in positive control samples (Figure S20).

##### Internal spike control present in culture positive and culture negative clinical samples

The internal spike of *Thermus thermophilus* was present in all culture positive and culture negative clinical samples. The median percentage of *T. thermophilus* reads was 3.6% (2-8) [0.7-58] in culture positive clinical samples (Figure S21), and 100% (100-100) [4.5-100] in culture negative samples (Figure S22).

#### Barcode contamination

Within each flow cell we estimated rates of mis-assigned reads (assigned to unused barcodes) and cross-assigned reads (assigned to incorrect in-use barcodes). The median probability for a read to be mis-assigned was 0.000054 (IQR, 0.000008-0.000129) [Range, 0-0.000379] for unfiltered reads and 0.000050 (0.000008-0.000119) [0-0.000372] for filtered reads, equating to 1 misassigned read per ~18,519 unfiltered and ~20,000 filtered reads (Figure S23). For cross-assigned reads, the median probability was 0.000028 (0.000019-0.000039) [0.000009-0.000199] (Figure S24), and in negative samples, it was 0.000021 (0.000010-0.000026) [0.000008-0.000052] (Figure S25). This translates to 1 cross-assigned read per ~35,714 in positive samples and ~47,619 in negative samples. Barcode 08 showed a significant association with off-target reads (p=0.004) (Figure S26).

#### Genes implicated in true and false AMR calls

ResFinder, using fastq reads at 24 hours, demonstrated superior performance in identifying resistance genes (Figure S12). The most frequently detected genes in true calls were *bla*TEM (n=324), *bla*Z (n=91), and *bla*CTX (n=90). However, *bla*TEM also caused the most false resistance (major error) calls, with 295 incorrect gene calls (IGC), followed by *bla*SHV (52x IGC), *bla*PAO (19x IGC), and others, particularly for ceftazidime, ceftriaxone, and piperacillin+tazobactam (Figure S13). Despite ResFinder's overall accuracy, AMRFinder and RGI (CARD) identified genes like *bla*EC and *bla*ACT that ResFinder missed, highlighting areas to reduce Very Major Errors (VMEs), while no tool was able to predict phenotypic resistance in 9 species-drug combinations (Table S9 and 10).
